# Sex differences in genetic and environmental influences on frailty and its relation to body mass index and education

**DOI:** 10.1101/2020.12.08.20245969

**Authors:** Jonathan K. L. Mak, Chandra A. Reynolds, Sara Hägg, Xia Li, Malin Ericsson, Nancy L. Pedersen, Juulia Jylhävä, Ralf Kuja-Halkola

## Abstract

**Background:** Frailty is a multifactorial expression of aging influenced by numerous genetic and environmental factors. However, sex differences in how these factors may affect frailty, and the gene-environment interplay among frailty and two of its well-established risk factors, unhealthy body mass index (BMI) and low education, are less clear.

**Methods:** In a large sample of 42,994 Swedish twins aged 41–99 years, we used structural equation models to estimate the genetic (heritability) and environmental sources of variance in frailty, defined as the Rockwood frailty index (FI), separately in men and women. We also estimated the genetic and environmental contributions to the associations between FI and BMI, and FI and education. Moderation models were fitted to test for gene-environment interaction by levels of BMI and education.

**Results:** Genetic and individual-specific environmental factors contributed approximately equally to the FI variance. The heritability of FI was slightly higher in women (52%) than in men (45%), yet we found only weak-to-no indication of different sources of genetic variance influencing frailty in men and women. A small-to-moderate genetic overlap between FI and BMI, and a perfect negative correlation of the environmental factors common to twins in a pair between FI and education were observed. Additionally, the heritability of FI was greater at both low and high BMI levels, with similar patterns of moderation in both sexes.

**Conclusions:** Individual differences in frailty are equally influenced by genetic and individual-specific environmental factors, and different mechanisms seem to underlie the association of frailty with BMI and education.

## Introduction

Frailty, as a manifestation of aging, is a state of increased vulnerability due to cumulative decline in multiple physiological systems (1). The concept of frailty has been operationalized in two principal models: the frailty phenotype (FP) defines frailty as a clinical syndrome associated with unintentional weight loss, weak grip strength, slow walking speed, exhaustion, and low physical activity (2), whereas the Rockwood frailty index (FI) summarizes vulnerability quantitatively and defines frailty as the accumulation of deficits from a wide range of physical and psychosocial functioning (3,4). Regardless of the definition being used, frailty has been consistently linked to adverse outcomes such as falls, hospitalizations, loss of independence, and mortality (5), posing a significant public health concern. Tackling frailty is also recognized as a priority by the European Union (6), calling for better understanding of the syndrome. The prevalence of frailty increases with age, with an overall estimated prevalence of 10.7% among those aged ≥65 and 26.1% among those aged ≥85, varying depending on which frailty scale is used (7). Meanwhile, there is sizable variability in the frailty status at all ages (8), highlighting the necessity to gain understanding on the underpinnings of the individual differences. The sex-paradox of frailty has been widely described, in which women experience higher levels of frailty but men are more vulnerable to death at any given level of frailty, yet the mechanisms behind remain elusive (9).

In accordance with its multifactorial nature, frailty is influenced by both genetic and environmental factors, with heritability estimates of the FI ranging from 30 to 45% (10,11). However, these studies included female twins only, leaving sex differences in the heritability of FI unaddressed. Recently, a large genome-wide association study (GWAS) revealed that the molecular genetic underpinnings of the FI are shared to some extent with key public health risk factors, such as body mass index (BMI), cardiovascular disease, smoking, as well as mental health traits (12). A vast body of literature has discerned the lifestyle and environmental determinants of frailty, highlighting the role of several physical, social, behavioral and psychological factors (13–15). There is also a persisting socioeconomic gradient in frailty, with increasing socioeconomic adversity associated with higher frailty and contributing to health inequalities even in old age (16). However, few studies have elucidated the mechanisms by which the risk factors influence frailty. For the two significant risk factors, unhealthy BMI ranges including both underweight and overweight (17–19) and low educational attainment (13,20–22), a recent Mendelian randomization analysis has shown that among all modifiable risk factors the genetic risk of higher BMI and lower educational attainment showed the strongest associations with the risk of frailty (12). Thus, there seems to be overlap in the genetic and/or environmental underpinnings of FI with BMI and education. Moreover, it is not known whether the genetic and environmental influences on the FI itself are altered by BMI and education levels, that is, whether there is gene-environment interaction (G×E) such that genetic risk of frailty may be amplified or suppressed with different environmental circumstances. It is also unclear whether men and women may differ with respect to G×E.

Twin studies offer a natural experiment to partition variance of traits into genetic and environmental etiologies by comparing the genetic similarities between monozygotic (MZ) and dizygotic (DZ) twins, who share 100% and ∼50% of their segregating alleles respectively. Large twin samples also allow robust assessment on whether the heritability of a trait differs quantitatively by sex, meaning whether it is the same genes but at different magnitudes that act upon the trait. Moreover, with the inclusion of opposite-sex twins, qualitative sex differences can be examined, informing whether the trait is influenced by different genetic factors in men and women. As the extension to classical twin models, bivariate models allow study of the sources of covariation between traits, while moderation models enable examination of G×E. To date, studies on the variance components of the FI are scarce and the potential sex differences remain unaddressed. The mechanisms by which high BMI and low education influence frailty are likewise poorly known. Not only is understanding the basis of the sex differences in frailty of importance, but identifying how the risk factors affect frailty will also aid in informing preventive strategies and public health interventions.

This study aimed to (i) provide a comprehensive estimation of the contribution of genetic and environmental factors to the variance of the FI, and identify potential quantitative and qualitative sex differences therein; (ii) explore the common genetic and environmental influences on the covariation of FI with BMI and education, two of the most prominent risk factors of frailty; and (iii) test for G×E by levels of BMI and education.

## Methods

### Study population

Twin participants were from the Screening Across the Lifespan Twin study (SALT), which is part of the population-based Swedish Twin Registry (23). During 1998–2002, all twins born in 1958 or before were invited to attend a telephone interview with questions on common diseases, symptoms, medication use, demographics, and lifestyle factors. With a response rate of 65% for those born in 1886–1925 and 74% for those born in 1926–1958, a total of 44,919 twin individuals participated in the survey (24). Zygosity was determined based on DNA, or questions on intra-pair similarities during childhood; the latter method was over 95% accurate when validated against DNA testing (25). This study has been approved by the Swedish Ethical Review Authority in Stockholm. Informed consent was obtained from all participants prior to data collection.

For the present analysis, we excluded those with uncertain zygosity and those who had over 20% missing data across the 44 frailty items, leaving *n*=42,994 consisting of 31,922 paired and 11,072 single respondents, aged from 41 to 99 years. Single twin individuals were retained as they contribute to the mean, variance and within-individual covariance estimates. Twins were grouped by zygosity: 4,788 MZ males (1,820 complete pairs), 5,997 MZ females (2,438 complete pairs), 7,640 DZ same-sex males (2,633 complete pairs), 8,808 DZ same-sex females (3,279 complete pairs), and 15,761 DZ opposite-sex individuals (5,791 complete pairs).

### Measures

We constructed an FI based on Rockwood’s deficit accumulation model (3), using a total of 44 self-reported frailty items selected from a wide range of health-status related symptoms, signs, disabilities, and diseases in various biological systems (**Supplementary Table 1**). Each respondent’s frailty items were summed up and divided by the total number of deficits measured (i.e., 44 in this study), yielding a continuous FI score ranging from 0 to 1. For instance, an individual with five deficits would have an FI of 5/44=0.11. The FI in SALT has previously been validated for its ability to predict mortality (26). Information on body mass index (BMI) and education were obtained from self-reported data. BMI was calculated as weight (kg) divided by height-squared (m^2^). We defined education as a continuous variable of the number of years of education completed.

### Statistical analysis

#### Twin design

The classical twin design allows decomposition of phenotypic variance into genetic and environmental sources. Genetic sources of variance include additive genetic variance (A, representing the sum of allelic effects at different loci that influence the trait) and dominance genetic variance (D, representing interactions between alleles at the same locus). In MZ twins, both A and D correlate 100%; while in DZ twins, A and D are assumed to correlate 50% and 25% respectively. The sum of A and D influences is referred to as the broad-sense heritability (H), which is the proportion of phenotypic variance that can be explained by genetic effects. On the other hand, environmental sources of variance include common environmental factors (C) such as family environment and environments that are in common or correlated in adulthood, which correlate 100% in both MZ and DZ twins; and unique environmental factors (E) which are uncorrelated between twins and contribute to differences between them. The E component also contains measurement error. In addition to variance, phenotypic covariance can also be partitioned into the same sources of influence, indicating the extent to which genetic and environmental variance components overlap across two or more traits.

#### Phenotypic, intraclass and cross-twin cross-trait correlations

For initial inspection of the familial similarity for MZ and DZ twin pairs, we estimated intraclass correlations for FI, BMI and education, which show how much each trait correlates within twin pairs. A higher MZ correlation than DZ correlation indicates contribution of genetics to variance in the trait. If DZ correlation is less than half of the MZ correlation, the D component is inferred; on the contrary, the C component is implied if DZ correlation is greater than half of the MZ correlation. A MZ correlation less than 1 indicates presence of E. Within-individual between-trait correlations, which we refer to as “phenotypic correlations”, were calculated for FI with BMI and education. To explore the genetic and environmental sources of covariances, we computed cross-twin cross-trait correlations, which represent the strength of relationship between one trait of the first twin and another trait of the second twin. They can be interpreted in a similar way as intraclass correlations. For example, a larger cross-twin cross-trait correlation in MZ than DZ twins suggests that phenotypic correlation between the two traits is partially owing to common genetic influences; and if MZ cross-twin cross-trait correlation is less than the phenotypic correlation then E is present. Phenotypic, intraclass and cross-twin cross-trait correlations were estimated separately across zygosity from constrained saturated models in which means, variances and phenotypic correlations were equated across twin order. Age was regressed out of the means (as linear effect for FI and linear+quadratic effect for BMI and education, after initial investigation of associations between variables), so that twin correlations are not spuriously inflated due to the same age of twins in pairs.

#### Univariate twin modelling

Before model fitting, we checked and observed no violations in the assumptions of equal means and variances across twin order and zygosity. Univariate twin models were fitted to estimate variance components of FI, after adjusting for age. Since C and D are confounded in classical twin modelling, either an ACE or ADE model can be fitted at a time. Sex differences were allowed in the models to examine whether genetic influence on FI differs across sex quantitatively (i.e., different magnitude of heritability in men and women) and qualitatively (i.e., different genetic sources in men and women). Quantitative sex differences were modelled by allowing variance components to differ among men and women. Qualitative sex differences were estimated by multiplying a genetic correlation term (*r*_fm_) to the expected genetic covariance of opposite-sex twin pairs, where a value of 1 would imply maximum genetic correlation (details on modelling qualitative sex difference are described in **Appendix Methods**). ACE or ADE sex-limitation models were then compared with their reduced models. We examined quantitative sex difference by constraining the broad-sense heritability to be equal in men and women (but allowing total variance to differ across sex) and tested if there was a significant reduction in model fit. Qualitative sex difference was assessed by testing whether *r*_fm_ was significantly less than 1. AE models were fitted to determine if the C and D parameters can be excluded from ACE and ADE models.

#### Bivariate twin modelling

During assumption testing, all means and variances could be equated across twin order and zygosity, except that equating means of education across zygosity led to a significant decrease in model fit so they were estimated separately in subsequent bivariate models. A “correlated factor model” (27) was applied to estimate the etiological correlations between genetic (*r*_A_ and *r*_D_) and environmental (*r*_C_ and *r*_E_) variance components of FI with BMI and education, apart from estimating variance components of each variables. We define the correlation between A+D in trait X and A+D in trait Y as the broad-sense genetic correlation, *r*_H_. Correlation of ±1 indicates complete overlap, while 0 indicates no overlap. Bivariate heritability was also calculated, defined as the proportion of phenotypic covariance explained by genetic covariation. Since estimates of A and D components are highly dependent as shown by non-significant A and D estimates, we estimated the broad-sense heritability and interpreted in ADE models to indicate the total genetic effects. Only same-sex twins were included in the bivariate quantitative sex-limitation models; variance components and etiological correlations were estimated separately by sex, after adjusting for age.

#### Moderation analysis

In previous univariate and bivariate models, we assumed that variance components of FI were constant across the population (i.e., no gene-environment interaction). We relaxed this assumption and examined whether the genetic and environmental influences on FI are moderated by different levels of BMI and education. Moderation may occur on the variance that is unique to FI; for example, changes in genetic variance components of FI with years of education would suggest differential genetic sensitivity of FI to education levels. Furthermore, moderation may exist on the covariance between FI and the moderator, for example, the genetic correlation between FI and BMI may vary with different levels of BMI. To investigate these possible moderating effects, we adopted the full bivariate moderation models developed by Purcell (28), and the extended univariate moderation models proposed by van der Sluis *et al* (29) (**Supplementary Figure 1–2**). By the full bivariate moderation models, we first assessed whether there were significant moderating effects on the covariance between FI and the moderators (BMI and education), on top of the variance unique to FI. In case of no statistically significant moderation on the covariance, the extended univariate moderation models were employed instead. All moderating effects were examined separately by sex in quantitative sex-limitation models using same-sex twins and controlled for age. Absolute genetic and environmental variance components of FI, as well as their proportions to the total variance, were plotted over BMI and education.

#### Sensitivity analysis

Since the FI variable is positively skewed (**Supplementary Figure 3**), we followed previous work and applied a square-root transformation to obtain an approximately normally-distributed FI (10,30). Univariate twin modelling and moderation analysis of the transformed FI were then performed to check for robustness of our results.

For all fitted models, we used the full information maximum-likelihood modelling in the R package OpenMx (version 2.17.3) for estimation of parameters that best fit the observed data. Changes in goodness of fit of models (distributed as χ^2^) were assessed by likelihood ratio tests, where significant values indicate worse fit of observed data. Akaike information criterion (AIC; a lower value is better) was used to select the most parsimonious and best-fitting models.

## Results

### Sample characteristics

**Table 1** shows the descriptive statistics of the sample, among the 42,994 twin individuals the mean age of which was 58.8 [standard deviation (SD) 10.7]. The median FI was 0.108 (interquartile range 0.062–0.176). Women constituted 53.6% of the sample; they were on average older (mean 59.2; SD 11.0) than men (mean 58.4; SD 10.4), and had a higher median FI (0.119 vs 0.097). Mean BMI and education were 25.0 kg/m^2^ (SD 3.5) and 10.5 years (SD 3.2) respectively; both were similar across sex.

**Table 1.** Characteristics of study population by sex

| Characteristic | Total<br>(n=42,994) | Men<br>(n=19,940) | Women<br>(n=23,054) |
| --- | --- | --- | --- |
| Age at interview, mean (SD) | 58.8 (10.7) | 58.4 (10.4) | 59.2 (11.0) |
| FI, median (IQR) | 0.108 (0.062, 0.176) | 0.097 (0.057, 0.153) | 0.119 (0.068, 0.193) |
| BMI (kg/m<sup>2</sup>), mean (SD) | 25.0 (3.5) | 25.5 (3.1) | 24.5 (3.8) |
| Education (years), mean (SD) | 10.5 (3.2) | 10.5 (3.2) | 10.4 (3.3) |
| Zygosity, n (%) |  |  |  |
| MZ | 10,785 (25.1) | 4,788 (24.0) | 5,997 (26.0) |
| *Twins from complete pairs* | 8,516 (79.0) | 3,640 (76.0) | 4,876 (81.3) |
| DZ same-sex | 16,448 (38.3) | 7,640 (38.3) | 8,808 (38.2) |
| *Twins from complete pairs* | 11,824 (71.9) | 5,266 (68.9) | 6,558 (74.5) |
| DZ opposite-sex | 15,761 (36.7) | 7,512 (37.7) | 8,249 (35.8) |
| *Twins from complete pairs* | 11,582 (73.5) | 5,791 (77.1) | 5,791 (70.2) |

*Note:* BMI, body mass index; DZ, dizygotic twins; FI, frailty index; IQR, interquartile range; MZ, monozygotic twins; SD, standard deviation

### Phenotypic, intraclass and cross-twin cross-trait correlations

Age-adjusted correlations across zygosity are presented in **Table 2** (unadjusted correlations in **Supplementary Table 2**). For FI, BMI and education, the intraclass correlations in MZ twins were larger than that in DZ twins and were <1, suggesting that both genetic and unique environmental factors influence on all three traits. We observed significantly greater intraclass correlations of FI in women than in men for same-sex twins, implying quantitative sex differences; nevertheless, intraclass correlation of FI in opposite-sex twins did not seem to be smaller than that in same-sex DZ twins, suggesting absence of qualitative sex differences. FI had a positive phenotypic correlation with BMI (*r*=0.13) and a negative correlation with education (*r=*-0.09). The higher MZ than DZ cross-twin cross-trait correlation between FI and BMI indicated that part of their covariation could be explained by genetic influences in common. In contrast, cross-twin cross-trait correlations between FI and education were comparable for MZ and DZ twins, indicating common environmental factors, rather than genetic factors, contributing to their covariance. All MZ cross-twin cross-trait correlations were less than the corresponding phenotypic correlations, suggesting unique environmental influence on the covariance between traits.

**Table 2.** Adjusted phenotypic correlations, intraclass correlations and cross-twin cross-trait correlations for the frailty index (FI), body mass index (BMI) and education

| Zygosity | Phenotypic correlations |  | Intraclass correlations |  |  | Cross-twin cross-trait correlations |  |
| --- | --- | --- | --- | --- | --- | --- | --- |
|  | FI & BMI | FI & Education | FI | BMI | Education | FI & BMI | FI & Education |
| Total | 0.13 (0.12, 0.14) | -0.09 (-0.10, -0.08) |  |  |  |  |  |
| MZ | 0.12 (0.10, 0.14) | -0.09 (-0.11, -0.07) | 0.51 (0.49, 0.53) | 0.68 (0.67, 0.70) | 0.65 (0.64, 0.67) | 0.10 (0.08, 0.12) | -0.07 (-0.10, -0.05) |
| DZ | 0.14 (0.13, 0.15) | -0.09 (-0.11, -0.07) | 0.18 (0.17, 0.20) | 0.23 (0.22, 0.25) | 0.42 (0.40, 0.43) | 0.06 (0.05, 0.07) | -0.08 (-0.09, -0.06) |
| MZ males | 0.14 (0.11, 0.17) | -0.10 (-0.13, -0.07) | 0.45 (0.41, 0.48) | 0.66 (0.63, 0.68) | 0.66 (0.63, 0.68) | 0.11 (0.08, 0.14) | -0.07 (-0.10, -0.04) |
| MZ females | 0.15 (0.12, 0.18) | -0.09 (-0.12, -0.06) | 0.53 (0.50, 0.56) | 0.69 (0.67, 0.71) | 0.65 (0.63, 0.67) | 0.13 (0.10, 0.16) | -0.08 (-0.11, -0.05) |
| DZ males | 0.12 (0.09, 0.14) | -0.09 (-0.11, -0.07) | 0.12 (0.09, 0.16) | 0.27 (0.24, 0.31) | 0.44 (0.42, 0.47) | 0.05 (0.02, 0.07) | -0.07 (-0.10, -0.05) |
| DZ females | 0.19 (0.17, 0.21) | -0.07 (-0.09, -0.05) | 0.24 (0.20, 0.27) | 0.31 (0.28, 0.34) | 0.46 (0.43, 0.48) | 0.10 (0.08, 0.13) | -0.06 (-0.09, -0.04) |
| DZ opposite-sex | 0.11 (0.09, 0.13) | -0.10 (-0.13, -0.08) | 0.17 (0.15, 0.20) | 0.18 (0.15, 0.20) | 0.38 (0.36, 0.41) | 0.03 (0.01, 0.06) | -0.09 (-0.11, -0.06) |

*Note:* MZ, monozygotic twins; DZ, dizygotic twins. Phenotypic correlations are the within-individual correlations between FI and BMI, and between FI and education. Intraclass correlations indicate the extent to which each trait correlates within twin pairs. Cross-twin cross-trait correlations show the extent to which FI of the first twin correlate with the other trait (i.e., BMI or education) of the second twin. 95% confidence intervals are presented in parentheses. All correlations were adjusted for age.

### Univariate twin modelling

Overall, the best-fitting univariate model of FI was an ADE model with only quantitative sex differences (**Table 3**). Compared to the saturated model (i.e., a model that fully describes the observed data), the full ADE model did not provide a worse model fit, indicating that the model fit the data well, but both the ACE and AE models provided a poor fit to the data. Removing quantitative sex differences from the ADE full sex-limitation model resulted in a reduced model fit; meanwhile, the *r*_fm_ was close to and statistically non-significantly different from unity, suggesting presence of quantitative, but not qualitative sex difference in the genetics of FI. We observed largely dissimilar proportions of additive and dominance genetic influences on FI for men and women. Alternatively, we estimated the broad-sense heritability of FI, which was 45% [95% confidence interval (CI) 41–48%] in men, and at a statistically significantly higher proportion of 52% (95% CI 50–55%) in women. The rest of the variation in FI was explained by unique environmental factors, which accounted for 55% (95% CI 52– 59%) and 48% (95% CI 45–50%) of the total variance in men and women, respectively.

**Table 3.** Model fitting results and parameter estimates from univariate sex-limitation models of the frailty index (FI)

| Model | Model fit statistics |  |  |  | Parameter estimates for men and women (95% CI) |  |  |  |  |
| --- | --- | --- | --- | --- | --- | --- | --- | --- | --- |
| | AIC | ΔLL | Δdf | $p$ | A | D/C | H | E | $r_{\text{fm}}$ |
| Saturated | 19953 | - | - | - | - | - | - | - | - |
| ADE full sex-limitation | 19940 | 19.1 | 16 | 0.264 | M: 7% (0, 23)<br>F: 41% (28, 55) | M: 38% (21, 55)<br>F: 11% (0, 25) | M: 44% (41, 48)<br>F: 52% (50, 55) | M: 56% (52, 59)<br>F: 48% (45, 50) | 0.69 (0.41, 0.96) |
| ADE qualitative sex-limitation | 19951 | 32.1 | 17 | 0.015 | M: 2% (0, 18)<br>F: 44% (31, 57) | M: 46% (31, 62)<br>F: 4% (0, 18) | M: 49% (47, 51)<br>F: 49% (47, 51) | M: 51% (49, 53)<br>F: 51% (49, 53) | 0.83 (0.15, 1.00) |
| **ADE quantitative sex-limitation** | **19939** | **19.7** | **17** | **0.288** | **M: 0% (0, 1)**<br>**F: 41% (28, 55)** | **M: 44% (41, 48)**<br>**F: 11% (0, 25)** | **M: 45% (41, 48)**<br>**F: 52% (50, 55)** | **M: 55% (52, 59)**<br>**F: 48% (45, 50)** | **1.00 (NA)** |
| ADE no sex difference | 19949 | 32.1 | 18 | 0.021 | M: 0% (0, 2)<br>F: 44% (31, 58) | M: 49% (45, 52)<br>F: 4% (0, 19) | M: 49% (47, 51)<br>F: 49% (47, 51) | M: 51% (49, 53)<br>F: 51% (49, 53) | 1.00 (NA) |
| ACE full sex-limitation | 19961 | 40.4 | 16 | 0.001 | M: 41% (37, 44)<br>F: 51% (49, 54) | M: 0% (0, 0)<br>F: 0% (0, 0) | M: 41% (37, 44)<br>F: 51% (49, 54) | M: 59% (56, 63)<br>F: 49% (46, 51) | 0.76 (0.64, 0.88) |
| AE full sex-limitation | 19957 | 40.4 | 18 | 0.002 | M: 41% (37, 44)<br>F: 51% (49, 54) | M: 0% (NA)<br>F: 0% (NA) | M: 41% (37, 44)<br>F: 51% (49, 54) | M: 59% (56, 63)<br>F: 49% (46, 51) | 0.76 (0.64, 0.88) |

*Note:* AIC, Akaike's Information Criterion; LL, log-likelihood; df, degrees of freedom; $p$ , $p$ -values of likelihood ratio tests compared with the saturated model. CIs are Wald-type confidence intervals with lower and upper bounds of 0 and 1 respectively. A, additive genetic factors; D, dominance genetic factors; C, common environmental factors; H, total genetic factors/ broad-sense heritability; E, unique environmental factors; $r_{\text{fm}}$ , genetic correlation between men and women, estimated using opposite-sex twins. M and F represents parameter estimates for men and women respectively. Full sex-limitation models allowed both quantitative and qualitative sex differences. In ADE qualitative sex-limitation model, broad-sense heritability of men and women were equated, but variance difference between sex was allowed. In ADE quantitative sex-limitation model, $r_{\text{fm}}$ was fixed to be 1. ACE and AE sub-models are not shown as the full models fit significantly worse than the saturated model. All models were adjusted for age. Best-fitting model is shown in bold.

### Bivariate twin modelling

The best-fitting bivariate model for FI and BMI was an ADE model (**Figure 1A**; model fit statistics and parameter estimates are listed in **Supplementary Tables 3–4**). Variance of BMI was predominantly explained by genetic factors, with broad-sense heritability estimated at 66% (95% CI 64–68%) in men and 69% (95% CI 67–71%) in women. We observed modest genetic correlations between FI and BMI in both men (*r*_H_=0.19; 95% CI 0.14–0.23) and women and (*r*_H_=0.26; 95% CI 0.22–0.29). The bivariate heritability for FI and BMI was 81% (95% CI 65–97%) and 87% (95% CI 78–95%) for men and women respectively, indicating that a substantial part of the correlation between FI and BMI could be attributable to genetic factors in common to both traits (**Figure 1C**). By contrast, the best-fitting bivariate model for FI and education according to AIC values was an ACE model (**Figure 1B**), although, all bivariate models between FI and education provided a worse fit than the saturated model. A moderate proportion of the variation in education was due to common environmental factors, which was 25% (95% CI 19–31%) in men and 29% (95% CI 23–34%) in women. The C component of FI was small and non-significant in both sexes, however, they had perfect negative correlations with that of education (*r*_C_=-1.00). Common environmental factors also accounted for considerable proportions of the phenotypic correlations between FI and education, which were 65% (95% CI: 23–107%) in men and 74% (95% CI: 22–126%) in women (**Figure 1C**). There was only weak overlap of genetic factors and unique environmental factors between FI and education, therefore, the FI-education correlation may primarily be phenotypic rather than shared at the underlying etiological level.

**Figure 1.**
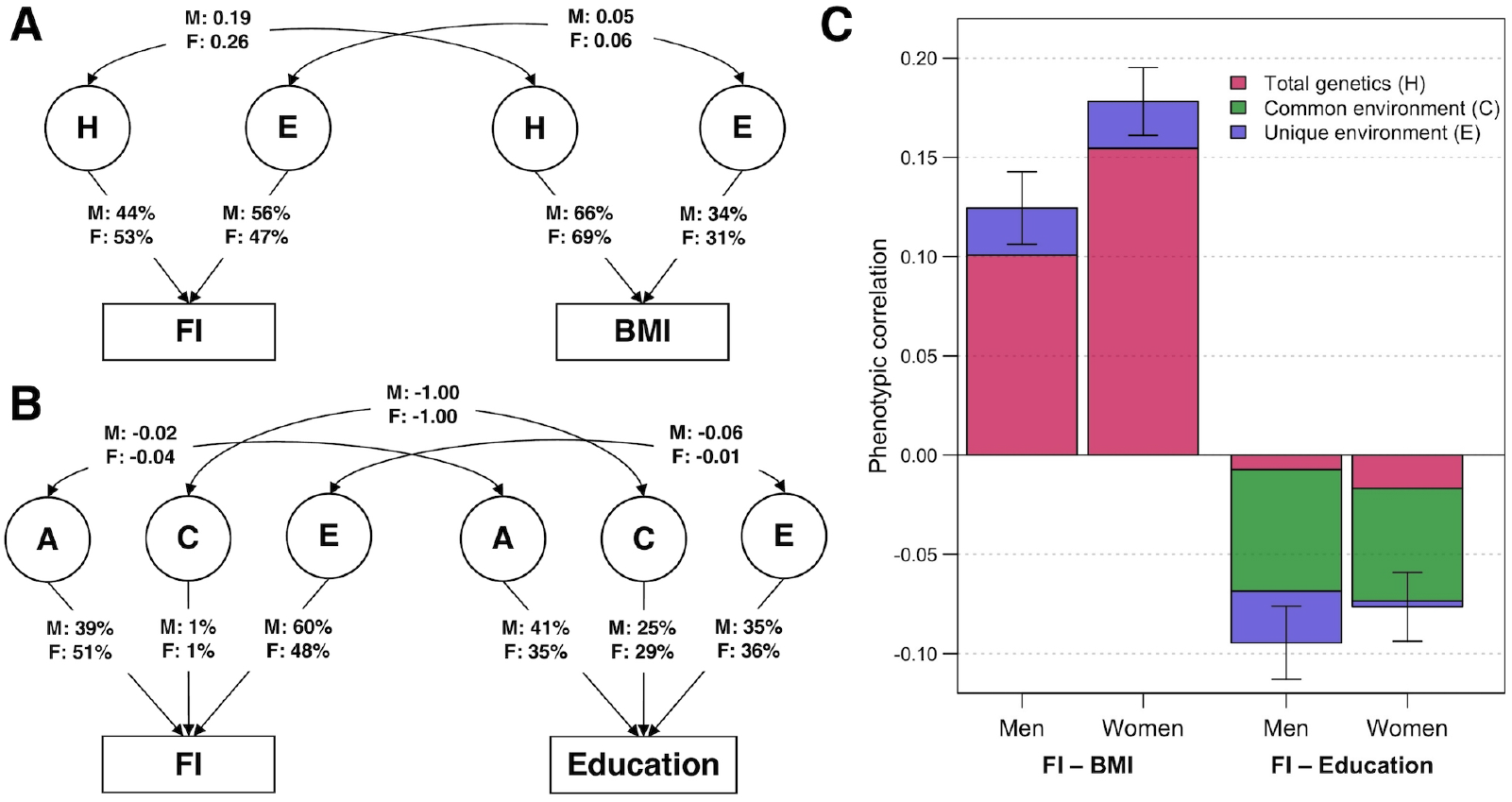
Parameter estimates from the best-fitting bivariate twin models. **(A)** ADE bivariate quantitative sex-limitation model between frailty index (FI) and body mass index (BMI), adjusted for age. Single headed-arrows represent the proportion of each traits explained by latent (circular) variance components; while double-headed arrows represent correlations between variance components. H indicates the sum of additive and dominance genetic factors; E indicates unique environmental factors; M and F are the estimates for men and women respectively. **(B)** ACE bivariate quantitative sex-limitation model between FI and education, adjusted for age. A indicates additive genetic factors; C indicates common environmental factors. **(C)** Phenotypic correlations of FI with BMI and education among men and women (with 95% confidence intervals), and the proportion of correlations explained by total genetic, common environmental and unique environmental factors. *Note:* Model-fitting results and parameter estimates can be found in Supplementary Tables 3–4.

### Moderation analysis

For moderation by BMI, we observed significant on the covariance between FI and BMI on top of the variance unique to FI; while for moderation by education, there was only significant moderating effects on the unique variance of FI (**Supplementary Tables 5–6**). Therefore, a full bivariate moderation model and an extended univariate moderation model were fitted for moderation by BMI and education, respectively. The H and E components of FI from the best-fitting moderation models are plotted in **Figure 2** (see **Supplementary Figure 4** for separate A and D components; and **Supplementary Figure 5** for separate common and unique variance estimates of FI by BMI levels). Patterns of moderation were consistent in both men and women, although women had a higher absolute variance of FI than men. Total variance of FI was larger at low and high BMI levels and at fewer years of education. Genetic factors accounted for a relatively larger proportions of the total FI variance for individuals with low and high BMI. In contrast, the relative proportions of genetic and environmental sources of FI variance did not seem to vary across education years.

**Figure 2.**
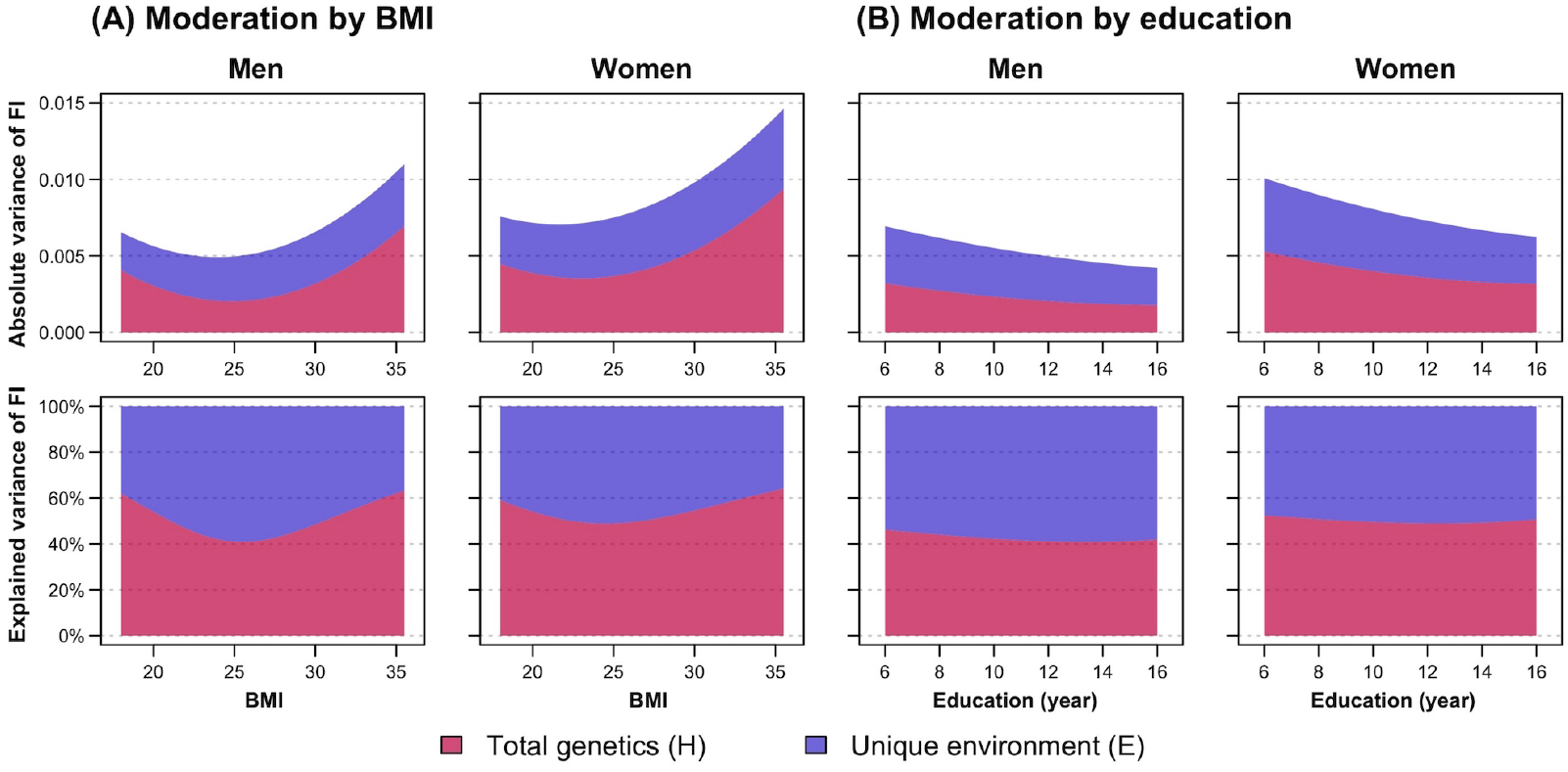
Moderation analysis of frailty index (FI) by **(A)** body mass index (BMI) and **(B)** education, stratified by sex. First row shows the absolute variance of FI, while the second row shows the proportion of FI variance explained by total genetic (H, indicating sum of additive and dominance genetic factors) and unique environmental (E) factors, with changes in BMI and education. Variance estimates of moderation by BMI were obtained from the full ADE bivariate moderation model between FI and BMI; while variance estimates of moderation by education were obtained from the ADE extended univariate moderation model between FI and education. Quantitative sex-differences were allowed in the models to obtain estimates separately for men and women. Models were adjusted for age. *Note:* Model-fitting results can be found in Supplementary Tables 5–6.

### Sensitivity analysis

Variance component estimates of the square-root transformed FI were largely consistent with that of the non-transformed FI variable (**Supplementary Table 7**). Moreover, similar patterns of moderation by BMI and education on the transformed FI were observed in men, but there was less evidence of moderation by BMI in women (**Supplementary Figure 6**),

## Discussion

Using a large sample of Swedish twins, we found that the variation in frailty was attributed rather equally to both genetic (with presence of additive and dominance effects) and individual-specific environmental influences. Heritability of FI was greater in women than in men, but there was no strong evidence of qualitative sex difference, indicating that the same genes are influencing FI in both sexes, although at different magnitudes. There was small-to-moderate genetic overlap between FI and BMI, and a complete overlap of the environmental factors common to twins within pairs between FI and education, suggesting different mechanisms on how BMI and education influence the risk of frailty. In addition, moderation analysis showed that the total variance of FI varies with levels of BMI and years of education, so that genetic influence on frailty tended to be greater at both low and high levels of BMI, but the heritability of FI did not seem to differ across education years.

Our finding that much of the FI variance is due to genetic factors is in line with two prior UK twin studies analyzing variance components of FI, which reported a heritability of 30–45% (10,11). By contrast, it is higher than the SNP-based heritability of 14% estimated in a recent GWAS (12), which may be partly due to the overestimation of heritability in twin studies (31), or that GWAS does not include non-additive genetic effects, as observed in the current study, and it has limited power to detect rare genetic variants that normally exist in complex traits like frailty (31,32). Environmental factors shared by members within twin pairs, which are usually those from childhood such as family environment, have negligible influence on frailty. Instead, a substantial proportion of the variation of FI is shaped by environmental factors unique to individuals, although this component also includes measurement error. This may reflect the multidimensional nature of frailty, in which diverse risk factors, such as underweight, obesity and low education, have been linked to the elevated risk of frailty (13–15).

To our knowledge, this is the first study that has formally examined sex differences in the heritability of frailty. We observed rather small, but significantly greater heritability in women (52%) than in men (45%), yet there was weak and statistically non-significant qualitative sex difference, indicating that it is probably the same sources of genetic variance affecting frailty in both sexes, but to different amounts. A higher heritability of some psychological and neurological traits, such as depression (33), pain (34), and insomnia (35), has also been observed among women, which is attributed at least partly to the higher prevalence of these traits in women. Similarly, women may also be more genetically susceptible to frailty, given that the genetic underpinnings of frailty has been found to be associated with neurological pathways (12). Another possible explanation to the difference in variance components across sex may be that men tend to report health problems less accurately (36), leading to an inflation of the E component. Notably, the total variance of FI was also higher in women than in men, reflecting the fact that women tend to have higher levels of frailty across the age range. The apparent sex difference may merely be owing to the overall greater variation of FI in women, instead of disparities in variance structure across sex, as shown in other traits such as BMI (37). Furthermore, the observed sex difference is not immense, yet statistically significant, perhaps also due to our large sample that provided enough statistical power. More research is warranted to further investigate if sex differences in genetic and environmental factors contribute to the sex-specificity of frailty.

Our second aim was to assess the overlap in the genetic and environmental influences of FI with BMI and education. There was a small-to-moderate genetic correlation between FI and BMI, suggesting that frailty and BMI may in part be influenced by the same genes. This may be attributable to their shared underlying mechanisms of energy metabolism and inflammation, known to be associated with both frailty (38,39) and BMI (40,41). Also, genes related to synaptic pathways were found to be enriched in the genetic architecture of frailty (12), and may likewise affect BMI (41). On the other hand, we found a relatively low genetic correlation between FI and education; instead, despite small and non-significant, the common environmental factors of FI had complete overlap with those of education, suggesting the relative importance of family environment on the education-frailty association. Previous studies found that genes associated with educational attainment have an inverse relationship with frailty (12,42); our findings may thus imply that education-associated genes are not directly influencing frailty, but indirectly through affecting individuals’ socioeconomic circumstances (43). The better family environment may then contribute to higher educational attainment and consequently prevention of frailty development, through the improved health literacy that higher education in the family brings (44,45), as well as the health seeking behavior that is especially characteristic to women with high education (46). It is however important to note that the common environmental factors of FI itself were small and not statistically significant and should be interpreted with caution.

Finally, we saw significant moderating effects on the genetic and unique environmental variance components of FI by both BMI and education, and the patterns were generally similar in men and women, although sensitivity analysis showed less evidence of moderation by BMI on the transformed FI in women. The overall variance of FI, as well as the relative proportion of genetic variance component were higher at low and high BMI levels, which follows the U-shape association between BMI and physical frailty reported in the literature (17–19). Therefore, in individuals whose BMI falls outside a healthy range, especially those with obesity (BMI>30), the increased risk of frailty may be due to their more pronounced expression of genetic susceptibilities to frailty. The reduced FI variance with increased years of education is congruent to previous literature showing smaller variance of health status at higher education levels (47). However, the proportions of genetic and unique environmental variance components over the total variance did not seem to differ over education years, suggesting that genetic and individual-specific environmental factors may have a stable contribution to frailty, independent of education.

This study included a population-representative adult twin sample, which provided enough statistical power to examine sex differences in heritability of frailty. We also used a validated FI as the frailty measure, which was shown to predict higher risks of all-cause and disease-specific mortality from midlife to old age (26). Nevertheless, several limitations should be considered. Firstly, given the cross-sectional nature of our data, we could not establish causal relationships among BMI, education and FI. Whether the variance components change with age should also be researched in future longitudinal studies with repeated measurements of frailty. Secondly, measures of frailty items, BMI, and education are all based on self-reported data, possibly causing misclassification and inaccurate reporting, especially in men (48). Thirdly, due to the different contribution of dominance genetic factors and common environmental factors to the variance of FI and education, all bivariate models for FI and education provided a poor fit, and we were unable to include both C and D parameters in the same statistical model with only reared-together twins in a classical twin design. Finally, there are some inherent limitations of twin modelling such as potential violation on the assumptions of equal environments (i.e., MZ and DZ twins are treated the same) and random mating in population (i.e., no assortment), although these should have minimal effects on the heritability estimates (49).

Overall, our findings demonstrate that individual differences in frailty are attributable to both genetic and individual-specific environmental factors. Sex differences are evident, in which women have a slightly higher heritability of frailty than men, but there is limited evidence of different genetic factors influencing frailty in men and women. Furthermore, the two main risk factors of frailty, BMI outside a healthy range and low education, seem to operate through different mechanisms in frailty development, highlighting the relative importance of genes and family environment on the associations of frailty with BMI and education, respectively. These results would help in expanding our current understanding on the individual differences in frailty.

## Data Availability

Data used in the current study are not publicly available due to ethical reasons. However, data are available upon request from the Swedish Twin Registry for researchers who meet the criteria for access to confidential data. Data from the SALT study are available from the Swedish Twin Registry steering committee (https://ki.se/en/research/the-swedish-twin-registry).

## Acknowledgements

C.A.R., S.H., J.J., and R.K.-H. contributed to the study design and statistical analysis plan. J.K.L.M., X.L., and R.K.-H. performed statistical analyses. N.L.P. is the leader of the SALT study and responsible for cohort recruitment, data collection, and funding. J.K.L.M., M.E., J.J., and R.K.-H. wrote the manuscript. All authors contributed to the interpretation of the results, and read and approved the final manuscript. We acknowledge the Swedish Twin Registry for access to data. The Swedish Twin Registry is managed by Karolinska Institutet and receives funding through the Swedish Research Council under the grant number 2017-00641.

