## Supplementary for "Sex differences in genetic and environmental influences on frailty and its relation to body mass index and education"

#### Supplementary Figures

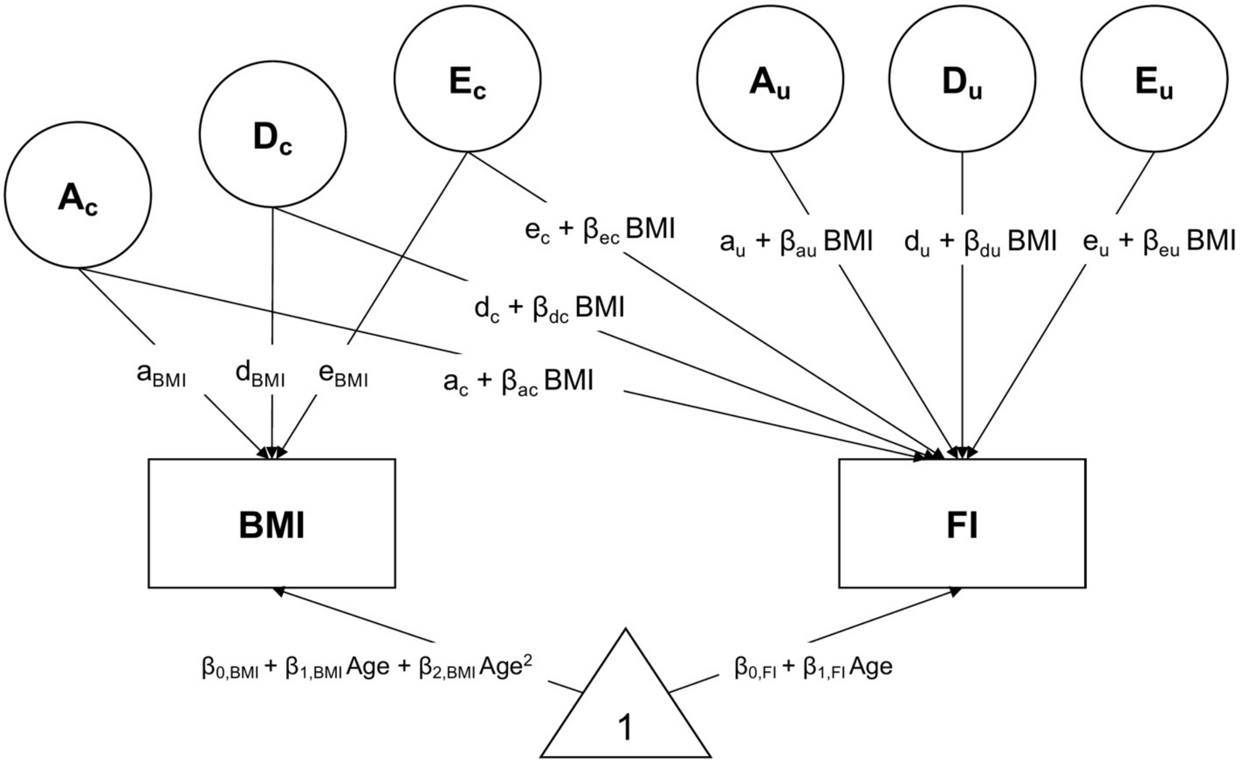

**Supplementary Figure 1.** Full bivariate moderation model between frailty index (FI) and body mass index (BMI) (for one twin). A, additive genetic factors; D, dominance genetic factors; E, unique environmental factors. $A_C$, $D_C$ and $E_C$ indicate genetic and environmental influences common to FI and BMI; while $A_U$, $D_U$ and $E_U$ indicate genetic and environmental influences unique to FI. Total variance of FI by BMI is the sum of common and unique variance estimates; for example, the total additive genetic variance components of FI by BMI can be calculated as: $(a_c + \beta_{ac}\ BMI)^2 + (a_u + \beta_{au}\ BMI)^2$.

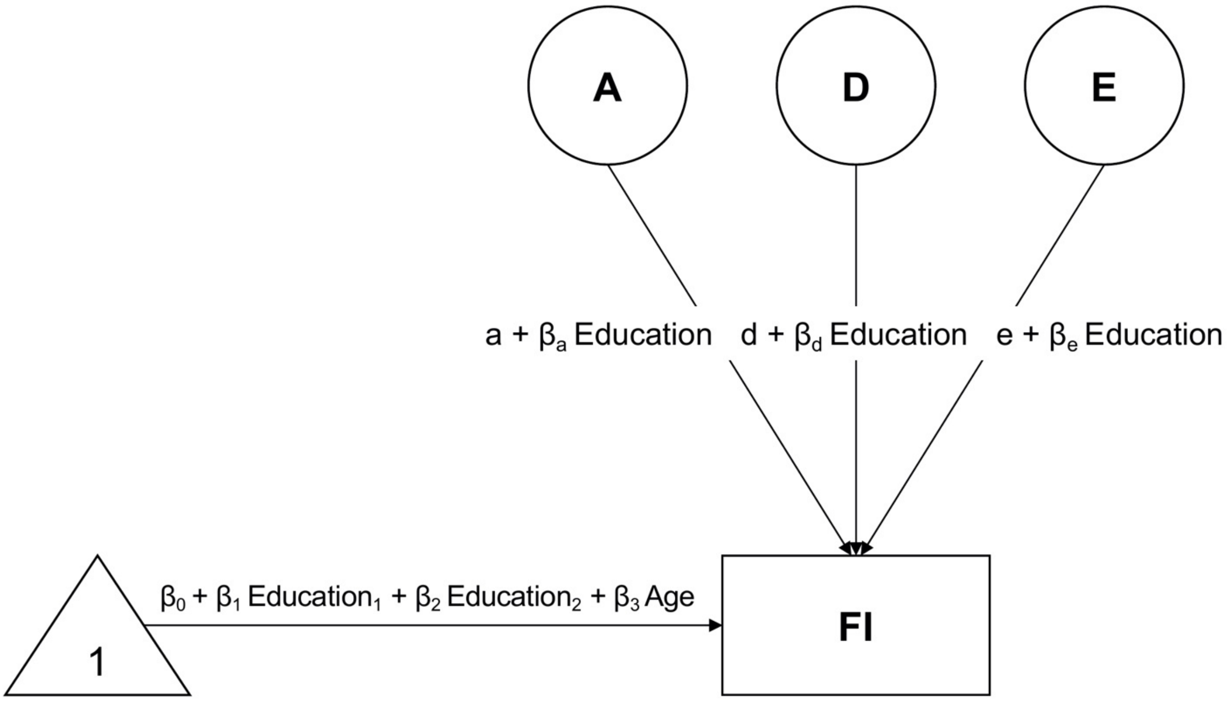

**Supplementary Figure 2.** Extended univariate moderation model between frailty index (FI) and education (for one twin). A, additive genetic factors; D, dominance genetic factors; E, unique environmental factors. Mean of FI is adjusted for the moderator (i.e. education) of both the individual and his/her co-twin.

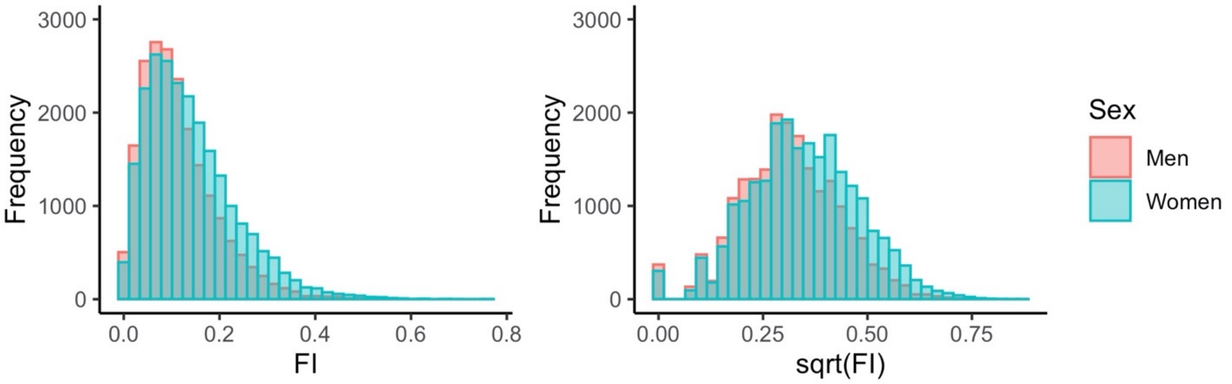

**Supplementary Figure 3.** Distribution of frailty index (FI) among men and women ($n$ = 42,994). Left panel shows the distribution of the untransformed FI, while right panel shows the distribution of the square-root transformed FI. Red color indicates men, while green color indicates women.

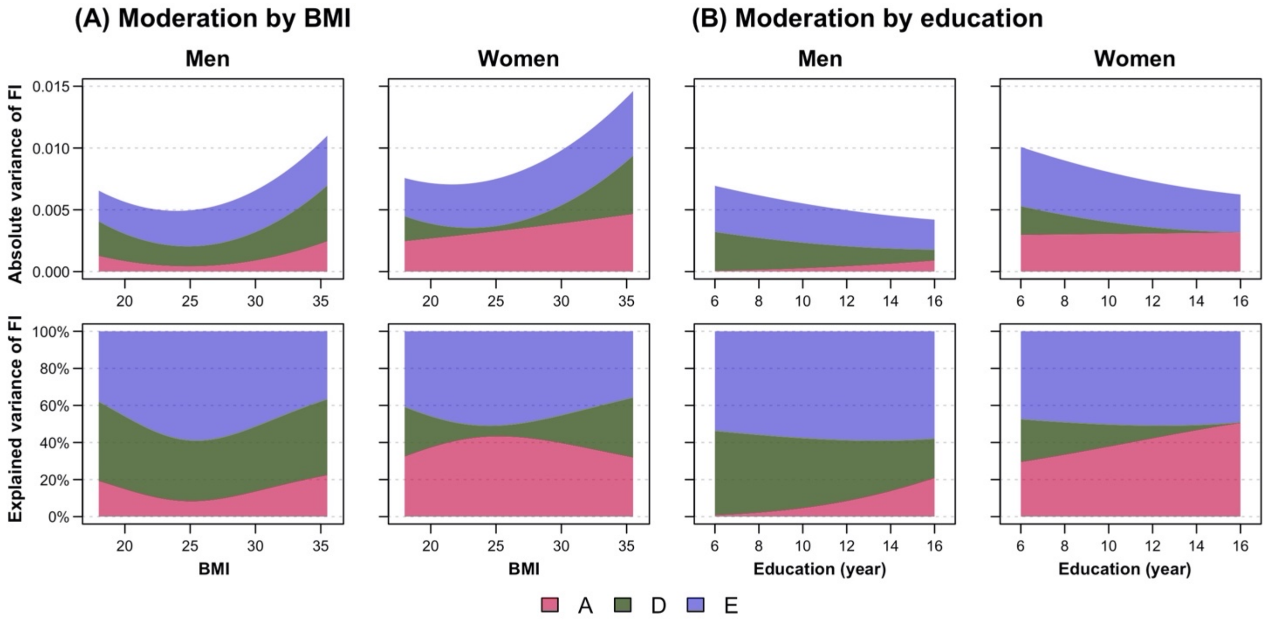

**Supplementary Figure 4.** Variance components of frailty index (FI) by **(A)** body mass index (BMI) and **(B)** education from moderation analysis, stratified by sex. First row shows the absolute variance of FI, while the second row shows the proportion of FI variance explained by additive genetic (A), dominance genetic (D) and unique environmental (E) factors, with changes in BMI and education. Variance estimates of moderation by BMI were obtained from the full ADE bivariate moderation model between FI and BMI; while the variance estimates of moderation by education were obtained from the ADE extended univariate moderation model between FI and education. Quantitative sex-differences were allowed in the models to obtain estimates separately for men and women. Models were adjusted for age.

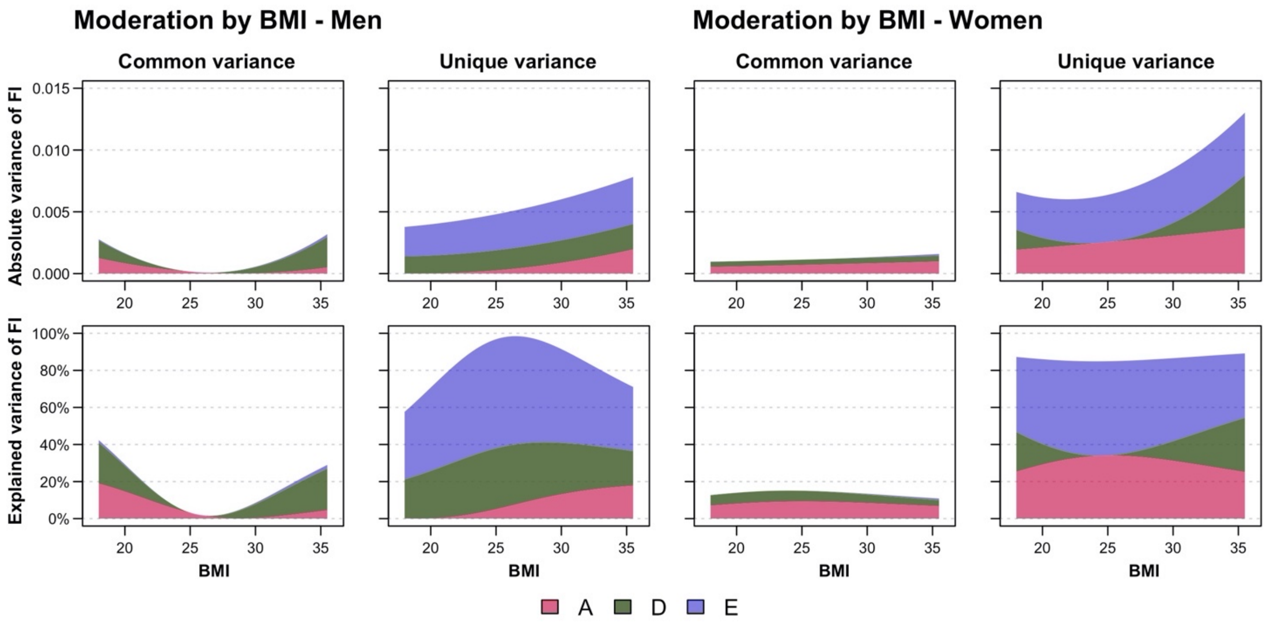

**Supplementary Figure 5.** Common and unique variance components of frailty index (FI) by body mass index (BMI) from moderation analysis, stratified by sex. First row shows the absolute variance of FI, while the second row shows the proportion of FI variance explained by additive genetic (A), dominance genetic (D) and unique environmental (E) factors, with changes in BMI levels. Variance estimates were obtained from the full ADE bivariate moderation model between FI and BMI. Quantitative sex-differences were allowed in the models to obtain estimates separately for men and women. Models were adjusted for age.

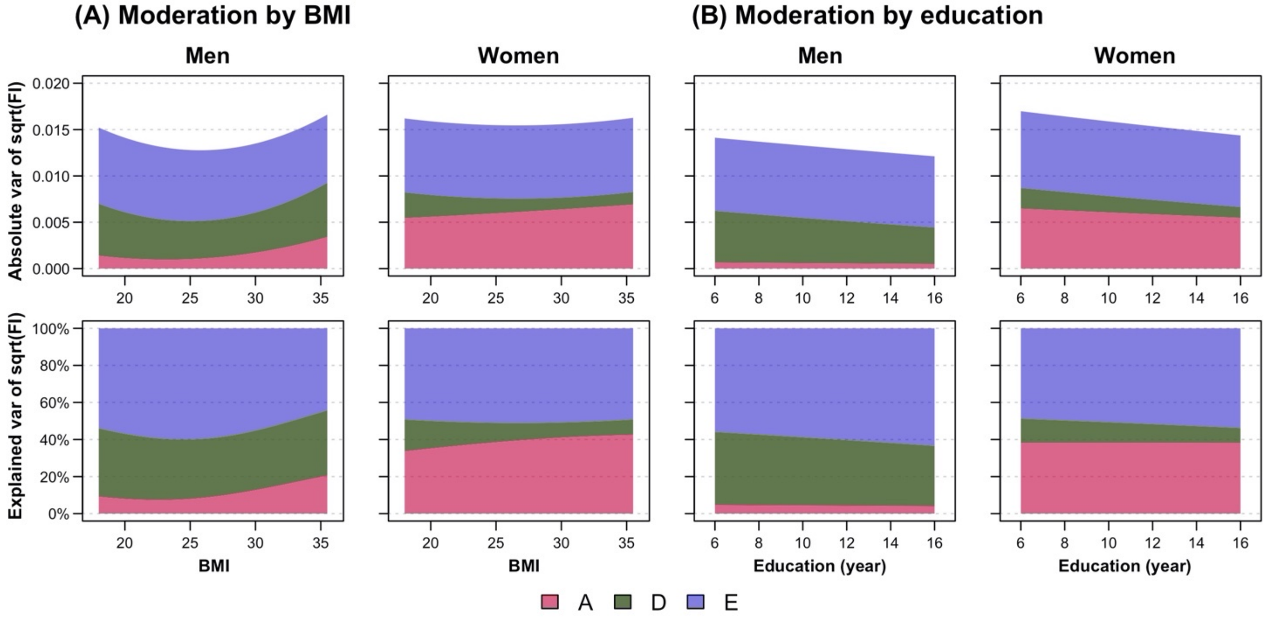

**Supplementary Figure 6.** Variance components of square-root transformed frailty index [sqrt(FI)] by **(A)** body mass index (BMI) and **(B)** education from moderation analysis, stratified by sex. First row shows the absolute variance of sqrt(FI), while the second row shows the proportion of sqrt(FI) variance explained by additive genetic (A), dominance genetic (D) and unique environmental (E) factors, with changes in BMI and education. Variance estimates of moderation by BMI were obtained from the full ADE bivariate moderation model between sqrt(FI) and BMI; while variance estimates of moderation by education were obtained from the ADE extended univariate moderation model between sqrt(FI) and education. Quantitative sex-differences were allowed in the models to obtain estimates separately for men and women. Models were adjusted for age.

#### Supplementary Tables

**Supplementary Table 1**. List of the 44 frailty items and the scoring used for construction of frailty index (FI)

| No. | Frailty item | Scoring |
|---|---|---|
| 1 | General health status | Excellent=0; Good=0.25; Average=0.5; Not so good=0.75; Bad=1 |
| 2 | Health status prevents from doing things normally would like to do | Not at all=0; To some extent=0.5; A great deal=1 |
| 3 | Serious infections per year (other than respiratory) | 0-1 time=0; 2-5 times=0.5; ≥5 times=1 |
| 4 | Buzzing in ears | No=0; One ear/ both ears=1 |
| 5 | Angina pectoris | No=0; Yes=1 |
| 6 | Heart attack | No=0; Yes=1 |
| 7 | Heart failure | No=0; Yes=1 |
| 8 | High blood pressure | No=0; Yes=1 |
| 9 | Lipid disorder (e.g. high cholesterol, high triglycerides) | No=0; Yes=1 |
| 10 | Vascular spasm in legs (intermittent claudication) | No=0; Yes=1 |
| 11 | Clot in leg (venous thrombosis) | No=0; Yes=1 |
| 12 | Cerebral hemorrhage or clot in brain (stroke) | No=0; Yes=1 |
| 13 | TIA attacks (temporary weakness, paralysis or reduction of sensibility) | No=0; Yes=1 |
| 14 | Irregular cardiac rhythm/ atrial fibrillation | No=0; Yes=1 |
| 15 | Chronic lung disease (incl. chronic bronchitis, emphysema) | No=0; Yes=1 |
| 16 | Dizziness | No=0; Yes=1 |
| 17 | Rheumatoid arthritis | No=0; Yes=1 |
| 18 | Knee joint problem | No=0; Yes=1 |
| 19 | Sciatica | No=0; Yes=1 |
| 20 | Osteoporosis | No=0; Yes=1 |
| 21 | Hip joint problem | No=0; Yes=1 |
| 22 | Back pain | No=0; Yes=1 |
| 23 | Neck pain | No=0; Yes=1 |
| 24 | Diabetes (incl. old age diabetes; excl. pregnancy diabetes) | No=0; Yes=1 |
| 25 | Goiter | No=0; Yes=1 |
| 26 | Glandular diseases (excl. goiter) | No=0; Yes=1 |
| 27 | Gall bladder problem | No=0; Yes=1 |
| 28 | Liver disease (e.g. cirrhosis) | No=0; Yes=1 |
| 29 | Gout | No=0; Yes=1 |
| 30 | Kidney disease | No=0; Yes=1 |
| 31 | Stomach or intestine problems | No=0; Yes=1 |
| 32 | Recurring urinary tract problems | No=0; Yes=1 |
| 33 | Cancer, tumor disease or leukemia | No=0; Yes=1 |
| 34 | Migraine | No=0; Yes=1 |
| 35 | Asthma | No=0; Yes=1 |
| 36 | Allergy | No=0; Yes=1 |
| 37 | Recurrent periods of coughing | No=0; Yes=1 |
| 38 | Feeling depressed during the past week | Never/ almost never=0; Seldom=0.5; Often/ always/ almost always=1 |
| 39 | Feeling happy during the past week | Never/ almost never=0; Seldom=0.5; Often/ always/ almost always=1 |
| 40 | Feeling lonely during the past week | Never/ almost never=0; Seldom=0.5; Often/ always/ almost always=1 |
| 41 | Physical handicap | No=0; Yes=1 |
| 42 | Crohn's disease or ulcerative colitis | No=0; Yes=1 |
| 43 | Vision | Good=0; Reduced=0.5; Highly reduced/ blind=1 |
| 44 | Hearing | Good=0; Reduced=0.5; Highly reduced=1 |

**Supplementary Table 2.** Unadjusted phenotypic correlations, intraclass correlations and cross-twin cross-trait correlations for frailty index (FI), body mass index (BMI) and education

| | Phenotypic correlations | | Intraclass correlations | | | Cross-twin cross-trait correlations | |
|---|---|---|---|---|---|---|---|
| Zygosity | FI & BMI | FI & Education | FI | BMI | Education | FI & BMI | FI & Education |
| Total | 0.13 (0.12, 0.14) | -0.17 (-0.18, -0.16) | | | | | |
| MZ | 0.12 (0.09, 0.14) | -0.17 (-0.19, -0.15) | 0.53 (0.51, 0.56) | 0.69 (0.67, 0.70) | 0.71 (0.69, 0.72) | 0.09 (0.07, 0.12) | -0.16 (-0.18, -0.13) |
| DZ | 0.14 (0.13, 0.15) | -0.17 (-0.18, -0.16) | 0.23 (0.21, 0.25) | 0.24 (0.22, 0.26) | 0.50 (0.49, 0.52) | 0.07 (0.06, 0.08) | -0.16 (-0.17, -0.15) |
| MZ males | 0.11 (0.08, 0.14) | -0.17 (-0.20, -0.14) | 0.47 (0.43, 0.51) | 0.66 (0.63, 0.68) | 0.69 (0.67, 0.72) | 0.09 (0.05, 0.12) | -0.14 (-0.17, -0.10) |
| MZ females | 0.16 (0.13, 0.19) | -0.18 (-0.21, -0.15) | 0.55 (0.52, 0.58) | 0.69 (0.67, 0.71) | 0.72 (0.70, 0.74) | 0.14 (0.11, 0.17) | -0.17 (-0.20, -0.14) |
| DZ males | 0.10 (0.07, 0.12) | -0.17 (-0.19, -0.15) | 0.19 (0.15, 0.23) | 0.27 (0.24, 0.31) | 0.50 (0.47, 0.53) | 0.03 (0.00, 0.06) | -0.15 (-0.18, -0.13) |
| DZ females | 0.20 (0.18, 0.22) | -0.17 (-0.19, -0.15) | 0.28 (0.25, 0.31) | 0.32 (0.29, 0.35) | 0.57 (0.55, 0.59) | 0.12 (0.09, 0.14) | -0.16 (-0.19, -0.14) |
| DZ opposite-sex | 0.10 (0.08, 0.13) | -0.17 (-0.19, -0.15) | 0.21 (0.18, 0.23) | 0.18 (0.16, 0.21) | 0.47 (0.45, 0.49) | 0.03 (0.00, 0.06) | -0.14 (-0.17, -0.12) |

*Note*: MZ, monozygotic twins; DZ, dizygotic twins. Phenotypic correlations are the within-individual correlations between FI and BMI, and between FI and education. Intraclass correlations indicate the extent to which each trait correlates within twin pairs. Cross-twin cross-trait correlations show the extent to which FI of the first twin correlate with the other trait (i.e. BMI or education) of the second twin. 95% confidence intervals are presented in parentheses.

**Supplementary Table 3.** Model fitting results from bivariate models of frailty index (FI) with body mass index (BMI) and education

| Model | -2LL | df | AIC | ΔLL | Δdf | *p* |
|---|---|---|---|---|---|---|
| Bivariate FI and BMI | | | | | | |
| Saturated | 203422.9 | 53568 | 96287 | - | - | - |
| ACE bivariate | | | | | | |
| Quantitative sex-limitation | 203503.7 | 53602 | 96300 | 80.8 | 34 | $1.12\times10^{-5}$ |
| No sex difference | 203548.6 | 53609 | 96331 | 125.6 | 41 | $1.60\times10^{-10}$ |
| **ADE bivariate** | | | | | | |
| **Quantitative sex-limitation** | **203459.9** | **53602** | **96256** | **36.9** | **34** | **0.335** |
| No sex difference | 203485.9 | 53605 | 96276 | 63.0 | 37 | $4.90\times10^{-3}$ |
| AE bivariate | | | | | | |
| Quantitative sex-limitation | 203503.7 | 53608 | 96288 | 80.8 | 40 | $1.43\times10^{-4}$ |
| No sex difference | 203548.6 | 53612 | 96325 | 125.6 | 44 | $8.73\times10^{-10}$ |
| Bivariate FI and education | | | | | | |
| Saturated | 198109.0 | 54150 | 89809 | - | - | - |
| **ACE bivariate** | | | | | | |
| **Quantitative sex-limitation** | **198175.0** | **54182** | **89811** | **65.9** | **32** | $\mathbf{3.84\times10^{-4}}$ |
| No sex difference | 198208.4 | 54189 | 89830 | 99.4 | 39 | $3.53\times10^{-7}$ |
| ADE bivariate | | | | | | |
| Quantitative sex-limitation | 198300.4 | 54182 | 89936 | 191.4 | 32 | $1.29\times10^{-24}$ |
| No sex difference | 198315.7 | 54185 | 89946 | 206.7 | 35 | $3.18\times10^{-26}$ |
| AE bivariate | | | | | | |
| Quantitative sex-limitation | 198328.3 | 54188 | 89952 | 219.3 | 38 | $2.39\times10^{-27}$ |
| No sex difference | 198360.8 | 54192 | 89977 | 251.8 | 42 | $1.02\times10^{-31}$ |

*Note*: AIC, Akaike's Information Criterion; LL, log-likelihood; df, degrees of freedom; *p*, *p*-values of likelihood ratio tests compared with the saturated models. Opposite-sex twins were excluded in the bivariate analyses. All bivariate models between FI and education had significant worse model fit than the saturated model, since an ADE model fits better for FI while an ACE model fits better for education. During assumption testing, equating means of education across zygosity resulted in a significantly worse fit of data compared to the saturated model; therefore, means of education were estimated separately across zygosity in bivariate models between FI and education. All models were adjusted for age (as linear effect for FI, and linear+quadratic effect for BMI and education). Best-fitting models are shown in bold.

**Supplementary Table 4.** Parameter estimates (95% CI) from the best-fitting bivariate models

| | Variance components | | | | Genetic and environmental correlations | | | | Bivariate heritability | | | |
|---|---|---|---|---|---|---|---|---|---|---|---|---|
| Model | A | D/C | H | E | $r_A$ | $r_D$ / $r_C$ | $r_H$ | $r_E$ | Bivariate A | Bivariate D/C | Bivariate H | Bivariate E |
| **ADE bivariate model between FI and BMI** | | | | | | | | | | | | |
| FI | M: 6% (0, 22) | M: 38% (21, 54) | M: 44% (40, 48) | F: 56% (52, 60) | | | | | | | | |
| | F: 42% (28, 55) | F: 11% (0, 25) | F: 53% (50, 55) | F: 47% (45, 50) | | | | | | | | |
| BMI | M: 41% (27, 55) | M: 25% (10, 39) | M: 66% (64, 68) | M: 34% (32, 36) | M: 0.61 (-0.25, 1.46) | M: 0.01 (-0.33, 0.36) | M: 0.19 (0.14, 0.23) | M: 0.05 (0.01, 0.10) | M: 78% (-3, 159) | M: 3% (-82, 88) | M: 81% (65, 97) | M: 19% (3, 35) |
| | F: 56% (42, 69) | F: 13% (0, 27) | F: 69% (67, 71) | F: 31% (29, 33) | F: 0.48 (0.29, 0.67) | F: -0.64 (-1.70, 0.42) | F: 0.26 (0.22, 0.29) | F: 0.06 (0.02, 0.10) | F: 130% (79, 181) | F: -43% (-96, 10) | F: 87% (78, 95) | F: 13% (5, 22) |
| **ACE bivariate model between FI and education** | | | | | | | | | | | | |
| FI | M: 39% (35, 43) | M: 1% (0, 3) | M: 39% (35, 43) | M: 60% (56, 63) | | | | | | | | |
| | F: 51% (48, 54) | F: 1% (0, 3) | F: 51% (48, 54) | F: 48% (46, 51) | | | | | | | | |
| Education | M: 41% (34, 48) | M: 25% (19, 31) | M: 41% (34, 48) | M: 35% (32, 37) | M: -0.02 (-0.15, 0.11) | M: -1.00 (-1.00, -1.00) | M: -0.02 (-0.15, 0.11) | M: -0.06 (-0.10, -0.01) | M: 8% (-46, 61) | M: 65% (23, 107) | M: 8% (-46, 61) | M: 28% (6, 49) |
| | F: 35% (29, 42) | F: 29% (23, 34) | F: 35% (29, 42) | F: 36% (34, 38) | F: -0.04 (-0.15, 0.07) | F: -1.00 (-1.00, -1.00) | F: -0.04 (-0.15, 0.07) | F: -0.01 (-0.05, 0.03) | F: 22% (-40, 84) | F: 74% (22, 126) | F: 22% (-40, 84) | F: 4% (-17, 25) |

*Note*: BMI, body mass index; FI, frailty index; CI, Wald-type confidence interval; A, additive genetic factors; D, dominance genetic factors; H, total genetic factors/ broad-sense heritability; C, common environmental factors; E, unique environmental factors; *r*, correlation between variance components. M and F represents parameter estimates for men and women respectively. Bivariate heritability is the proportion of phenotypic correlation explained by genetic and environmental factors.

**Supplementary Table 5.** Model fitting results from moderation models of frailty index (FI) by body mass index (BMI)

| Model | -2LL | df | AIC | Comp | ΔLL | Δdf | *p* |
|---|---|---|---|---|---|---|---|
| ACE bivariate | | | | | | | |
| 1. Full moderation | 146418.0 | 38963 | 68492 | - | - | - | - |
| **ADE bivariate** | | | | | | | |
| **2. Full moderation** | **146354.9** | **38963** | **68429** | **-** | **-** | **-** | **-** |
| 3. Drop all covariance moderation | 146453.0 | 38969 | 68515 | 2 | 98.1 | 6 | $6.29\times10^{-19}$ |
| 4. Drop all moderation | 146653.5 | 38975 | 68704 | 2 | 298.7 | 12 | $9.01\times10^{-57}$ |
| AE bivariate | | | | | | | |
| 5. Full moderation | 146489.1 | 38973 | 68543 | 2 | 134.2 | 10 | $6.34\times10^{-24}$ |
| 6. Drop all covariance moderation | 146615.8 | 38977 | 68662 | 5 | 126.7 | 4 | $1.96\times10^{-26}$ |
| 7. Drop all moderation | 146697.7 | 38981 | 68736 | 5 | 208.6 | 8 | $9.73\times10^{-41}$ |

*Note*: AIC, Akaike's Information Criterion; Comp, model of comparison; df, degrees of freedom; LL, Log-likelihood; *p*, *p*-values of likelihood ratio tests compared with the models of comparison. Opposite-sex twins were excluded in the models. Quantitative sex differences were allowed to obtain separate estimates for men and women. All models were adjusted for age (as linear effect for FI, and linear+quadratic effect for BMI). Best-fitting model is shown in bold.

**Supplementary Table 6.** Model fitting results from moderation models of frailty index (FI) by education

| Models | -2LL | df | AIC | Comp | ΔLL | Δdf | *p* |
|---|---|---|---|---|---|---|---|
| ACE bivariate | | | | | | | |
| 1. Full moderation | 145282.4 | 40231 | 64820 | - | - | - | - |
| 2. Drop all covariance moderation | 145289.7 | 40237 | 64816 | 1 | 7.3 | 6 | 0.297 |
| 3. Drop all moderation | 145537.7 | 40243 | 65052 | 1 | 255.3 | 12 | $1.09{\times}10^{-47}$ |
| ADE bivariate | | | | | | | |
| 4. Full moderation | 145418.6 | 40231 | 64957 | - | - | - | - |
| 5. Drop all covariance moderation | 145423.2 | 40237 | 64949 | 4 | 4.6 | 6 | 0.592 |
| 6. Drop all moderation | 145665.6 | 40243 | 40243 | 4 | 247.0 | 12 | $5.71{\times}10^{-46}$ |
| AE bivariate | | | | | | | |
| 7. Full moderation | 145450.9 | 40241 | 64969 | 1 | 168.4 | 10 | $5.92{\times}10^{-31}$ |
| 8. Drop all covariance moderation | 145454.7 | 40245 | 64965 | 7 | 3.8 | 4 | 0.432 |
| 9. Drop all moderation | 145694.2 | 40249 | 65196 | 7 | 243.3 | 8 | $4.42{\times}10^{-48}$ |
| ACE extended univariate | | | | | | | |
| 10. Full moderation | 47816.3 | 20109 | 7598 | - | - | - | - |
| 11. Drop all moderation | 48064.9 | 20115 | 7835 | 10 | 248.6 | 6 | $8.24{\times}10^{-51}$ |
| **ADE extended univariate** | | | | | | | |
| **12. Full moderation** | **47793.4** | **20109** | **7575** | **-** | **-** | **-** | **-** |
| 13. Drop all moderation | 48039.4 | 20115 | 7809 | 12 | 246.0 | 6 | $2.90{\times}10^{-50}$ |
| AE extended univariate | | | | | | | |
| 14. Full moderation | 47823.0 | 20113 | 7597 | 12 | 29.6 | - | - |
| 15. Drop all moderation | 48064.9 | 20117 | 7831 | 14 | 241.9 | 4 | $3.69{\times}10^{-51}$ |

*Note*: AIC, Akaike's Information Criterion; Comp, model of comparison; df, degrees of freedom; LL, log-likelihood; *p*, *p*-values of likelihood ratio tests compared with the models of comparison. Opposite-sex twins were excluded in the models. Quantitative sex differences were allowed to obtain separate estimates for men and women. Bivariate models were adjusted for age (as linear effect for FI, and linear+quadratic effect for education); extended univariate models were adjusted for age, as well as education for both the individual and the co-twin. Best-fitting model is shown in bold. Due to the non-significant moderation on the covariance between FI and education in the bivariate models, the more parsimonious ADE extended univariate model was used.

**Supplementary Table 7.** Model fitting results and parameter estimates from univariate sex-limitation models of the square-root transformed frailty index [sqrt(FI)]

| | Model fit statistics | | | | Parameter estimates for men and women (95% CI) | | | | |
|---|---|---|---|---|---|---|---|---|---|
| Model | AIC | ΔLL | Δdf | $p$ | A | D/C | H | E | $r_{fm}$ |
| Saturated | -146609 | - | - | - | - | - | - | - | - |
| ADE full sex-limitation | -146620 | 20.6 | 16 | 0.196 | M: 7% (0, 23) | M: 34% (17, 50) | M: 41% (38, 45) | M: 59% (55, 62) | 0.68 (0.49, 0.93) |
| | | | | | F: 40% (27, 53) | F: 10% (0, 24) | F: 50% (47, 53) | F: 50% (47, 53) | |
| ADE qualitative sex-limitation | -146607 | 35.3 | 17 | 0.006 | M: 3% (0, 18) | M: 43% (27, 59) | M: 46% (44, 48) | M: 54% (52, 56) | 0.84 (0.18, 1.00) |
| | | | | | F: 43% (30, 56) | F: 3% (0, 17) | F: 46% (44, 48) | F: 54% (52, 56) | |
| **ADE quantitative sex-limitation** | **-146621** | **21.4** | **17** | **0.208** | **M: 0% (0, 1)** | **M: 41% (38, 45)** | **M: 42% (38, 45)** | **M: 58% (55, 62)** | **1.00 (NA)** |
| | | | | | **F: 40% (26, 53)** | **F: 10% (0, 24)** | **F: 50% (47, 53)** | **F: 50% (47, 53)** | |
| ADE no sex difference | -146609 | 35.2 | 18 | 0.009 | M: 0% (0, 3) | M: 46% (41, 50) | M: 46% (44, 48) | M: 54% (52, 56) | 1.00 (NA) |
| | | | | | F: 43% (29, 57) | F: 3% (0, 17) | F: 46% (44, 48) | F: 54% (52, 56) | |
| ACE full sex-limitation | -146602 | 38.7 | 16 | 0.001 | M: 38% (34, 41) | M: 0% (0, 0) | M: 38% (34, 41) | M: 62% (59, 66) | 0.77 (0.65, 0.90) |
| | | | | | F: 49% (46, 52) | F: 0% (0, 0) | F: 49% (46, 52) | F: 51% (48, 54) | |
| AE full sex-limitation | -146606 | 38.7 | 18 | 0.003 | M: 38% (34, 41) | M: 0% (NA) | M: 38% (34, 41) | M: 62% (59, 66) | 0.77 (0.65, 0.90) |
| | | | | | F: 49% (46, 52) | F: 0% (NA) | F: 49% (46, 52) | F: 51% (48, 54) | |

*Note*: AIC, Akaike's Information Criterion; LL, log-likelihood; df, degrees of freedom; $p$, $p$-values of likelihood ratio tests compared with the saturated model. CIs are Wald-type confidence intervals with lower and upper bounds of 0 and 1. A, additive genetic factors; D, dominance genetic factors; C, common environmental factors; H, total genetic factors/ broad-sense heritability; E, unique environmental factors; $r_{fm}$, genetic correlation between men and women, estimated using opposite-sex twins. M and F represents parameter estimates for men and women respectively. Full sex-limitation models allowed both quantitative and qualitative sex differences. In ADE qualitative sex-limitation model, broad-sense heritability of men and women were equated, but variance difference between sex was allowed. In ADE quantitative sex-limitation model, $r_{fm}$ was fixed to be 1. ACE and AE sub-models are not shown as the full models fit significantly worse than the saturated model. All models were adjusted for age. Best-fitting model is shown in bold.

### Appendix Methods

#### Introduction

In this appendix we outline an approach to estimating the genetic correlation between males and females for a phenotype which has genetic contributions both from additive and dominance deviations to its variance. *1.* Using the biometric model in a simplified two-allelic locus with equal allele frequencies, we will derive the resulting correlation between genetic contributions in males and females with different additive and dominance deviation contributions genotypic expression. *2.* We then show that the resulting correlation depends on allele frequencies. *3.* Finally, we suggest an intuitive solution for estimation of the genetic correlation between males and females, and compare its performance with other possible solutions in a series of simulations.

As outlined below, our suggested model for the covariance between twins in opposite sexed DZ pairs is

$$Cov\left(\begin{bmatrix} Y_1 \\ Y_2 \end{bmatrix}\right)_{osDZ} = \begin{bmatrix} \sigma_{Af}^2 + \sigma_{Df}^2 + \sigma_{Ef}^2 & r_{fm}\left(0.5\sigma_{Af}\sigma_{Am} + 0.25\sigma_{Df}\sigma_{Dm} + 0.25\sigma_{Af}\sigma_{Dm} + 0.25\sigma_{Df}\sigma_{Am}\right) \\ r_{fm}\left(0.5\sigma_{Af}\sigma_{Am} + 0.25\sigma_{Df}\sigma_{Dm} + 0.25\sigma_{Af}\sigma_{Dm} + 0.25\sigma_{Df}\sigma_{Am}\right) & \sigma_{Am}^2 + \sigma_{Dm}^2 + \sigma_{Em}^2 \end{bmatrix} \quad (1)$$

where $\sigma_X^2$ represent contributions to variance and covariance from source $X$. $X$ are $A$, additive genetic contributions, $D$, dominant genetic contributions, and $E$, individually unique contributions. The sub-indexes are also complemented with $f$ and $m$ to indicate female and male sources. Below we show that this model, although not uniformly unbiased, has some features which makes it suitable for situations where additive and dominance contributions to variance of the phenotype exists in both sexes (possibly in different proportions).

##### 1. Derivation of correlation due to genetics from biometric model

Following the set-up of the biometric model in Neale and Maes [1] we define one autosomal locus having allele $A$ and $a$ at equal ½ frequencies in a population. We define the genotypic effect on $Y$ to be $-d$ for allele combination $aa$, $h$ for allele combination $Aa$, and $d$ for allele combination $AA$ (see **Supplementary Figure 7**). Note that the definition of genotypic effect is somewhat arbitrary, see e.g. Neale and Maes [1].

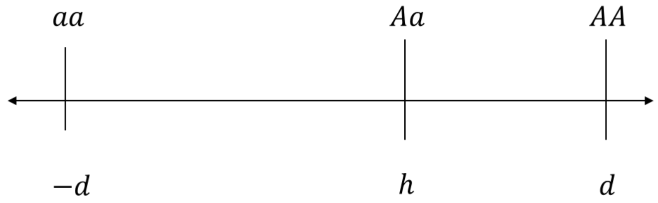

**Supplementary Figure 7.** Graphical representation of the genotypic effect. (Adapted from Neale and Maes [1])

Here $h$ indicates the deviation from additivity; we assume $h$ to be bounded between $-d$ and $d$ to keep a biologically feasible interpretation of additivity *vs* dominance/recessiveness. The ½ frequencies means that a random individual has a ¼ probability of having allele combination $aa$, a ½ probability of having $Aa$, and a ¼ probability of having $AA$. The mean genotypic contribution can be written as (using the law of total expectation)

$$\mu_Y = E(Y) = E(Y|aa)\Pr(aa) + E(Y|Aa)\Pr(Aa) + E(Y|AA)\Pr(AA) = -d \cdot \frac{1}{4} + h \cdot \frac{1}{2} + d \cdot \frac{1}{4} = \frac{1}{2}h \qquad (2)$$

The variance can be calculated as

$$\begin{aligned} Var(Y) &= E\left(\left(Y - E(Y)\right)^2\right) = E(Y^2) - E(Y)^2 \\ &= E(Y^2|aa)\Pr(aa) + E(Y^2|Aa)\Pr(Aa) + E(Y^2|AA)\Pr(AA) - \left(\frac{1}{2}h\right)^2 \\ &= (-d)^2 \cdot \frac{1}{4} + h^2 \cdot \frac{1}{2} + d^2 \cdot \frac{1}{4} - \frac{1}{4}h^2 = \frac{1}{2}d^2 + \frac{1}{4}h^2 \end{aligned} \qquad (3)$$

Now, let the same locus contribute to two phenotypes, $Y_1$ and $Y_2$ (e.g., same phenotype in males and females), possibly with different additivity and dominance deviation. Define the genotypic effects to be $\{-d_1, h_1, d_1\}$ and $\{-d_2, h_2, d_2\}$ for $Y_1$ and $Y_2$, respectively. Using the frequencies of allele combination in full siblings (or DZ twins), following Neale and Maes [1], we can create a table indicating the contributions to the covariance from each combination of allele in the two siblings.

**Supplementary Table 8.** Contributions to covariance, and expected frequency, between two DZ twins

| Sibling 1 alleles | Sibling 2 alleles | Genotypic effect minus mean, sibling 1 | Genotypic effect minus mean, sibling 2 | Contribution to covariance | Frequency |
|---|---|---|---|---|---|
| $AA$ | $AA$ | $d_1 - \frac{1}{2}h_1$ | $d_2 - \frac{1}{2}h_2$ | $d_1d_2 - \frac{1}{2}d_1h_2 - \frac{1}{2}h_1d_2 + \frac{1}{4}h_1h_2$ | $\frac{9}{64}$ |
| $AA$ | $Aa$ | $d_1 - \frac{1}{2}h_1$ | $\frac{1}{2}h_2$ | $\frac{1}{2}d_1h_2 - \frac{1}{4}h_1h_2$ | $\frac{6}{64}$ |
| $AA$ | $aa$ | $d_1 - \frac{1}{2}h_1$ | $-d_2 - \frac{1}{2}h_2$ | $-d_1d_2 - \frac{1}{2}d_1h_2 + \frac{1}{2}h_1d_2 + \frac{1}{4}h_1h_2$ | $\frac{1}{64}$ |
| $Aa$ | $AA$ | $\frac{1}{2}h_1$ | $d_2 - \frac{1}{2}h_2$ | $\frac{1}{2}h_1d_2 - \frac{1}{4}h_1h_2$ | $\frac{6}{64}$ |
| $Aa$ | $Aa$ | $\frac{1}{2}h_1$ | $\frac{1}{2}h_2$ | $\frac{1}{4}h_1h_2$ | $\frac{20}{64}$ |
| $Aa$ | $aa$ | $\frac{1}{2}h_1$ | $-d_2 - \frac{1}{2}h_2$ | $-\frac{1}{2}h_1d_2 - \frac{1}{4}h_1h_2$ | $\frac{6}{64}$ |
| $aa$ | $AA$ | $-d_1 - \frac{1}{2}h_1$ | $d_2 - \frac{1}{2}h_2$ | $-d_1d_2 + \frac{1}{2}d_1h_2 - \frac{1}{2}h_1d_2 + \frac{1}{4}h_1h_2$ | $\frac{1}{64}$ |
| $aa$ | $Aa$ | $-d_1 - \frac{1}{2}h_1$ | $\frac{1}{2}h_2$ | $-\frac{1}{2}d_1h_2 - \frac{1}{4}h_1h_2$ | $\frac{6}{64}$ |
| $aa$ | $aa$ | $-d_1 - \frac{1}{2}h_1$ | $-d_2 - \frac{1}{2}h_2$ | $d_1d_2 + \frac{1}{2}d_1h_2 + \frac{1}{2}h_1d_2 + \frac{1}{4}h_1h_2$ | $\frac{9}{64}$ |

We can thus calculate the contribution to covariance from this locus between phenotypes and between DZ twins as

$$
\begin{aligned}
Cov(Y_1, Y_2) &= \sum E\left((Y_1 - \mu_{Y_1})(Y_2 - \mu_{Y_2})\right) \\
&= \sum E\left((Y_1 - \mu_{Y_1})(Y_2 - \mu_{Y_2})|\text{allelic combination}\right) \Pr(\text{allelic combination}) = \\
&= \frac{9}{64}\left(d_1 d_2 - \frac{1}{2} d_1 h_2 - \frac{1}{2} h_1 d_2 + \frac{1}{4} h_1 h_2\right) + \frac{6}{64}\left(\frac{1}{2} d_1 h_2 - \frac{1}{4} h_1 h_2\right) \\
&+ \frac{1}{64}\left(-d_1 d_2 - \frac{1}{2} d_1 h_2 + \frac{1}{2} h_1 d_2 + \frac{1}{4} h_1 h_2\right) + \frac{6}{64}\left(\frac{1}{2} h_1 d_2 - \frac{1}{4} h_1 h_2\right) + \frac{20}{64}\left(\frac{1}{4} h_1 h_2\right) \\
&+ \frac{6}{64}\left(-\frac{1}{2} h_1 d_2 - \frac{1}{4} h_1 h_2\right) + \frac{1}{64}\left(-d_1 d_2 + \frac{1}{2} d_1 h_2 - \frac{1}{2} h_1 d_2 + \frac{1}{4} h_1 h_2\right) \\
&+ \frac{6}{64}\left(-\frac{1}{2} d_1 h_2 - \frac{1}{4} h_1 h_2\right) + \frac{9}{64}\left(d_1 d_2 + \frac{1}{2} d_1 h_2 + \frac{1}{2} h_1 d_2 + \frac{1}{4} h_1 h_2\right) \\
&= \frac{1}{64}(9 - 1 - 1 + 9) d_1 d_2 + \frac{1}{64}\left(-\frac{9}{2} + \frac{6}{2} - \frac{1}{2} + \frac{1}{2} - \frac{6}{2} + \frac{9}{2}\right) d_1 h_2 \\
&+ \frac{1}{64}\left(-\frac{9}{2} + \frac{1}{2} + \frac{6}{2} - \frac{6}{2} - \frac{1}{2} + \frac{9}{2}\right) h_1 d_2 \\
&+ \frac{1}{64}\left(\frac{9}{4} - \frac{6}{4} + \frac{1}{4} - \frac{6}{4} + \frac{20}{4} - \frac{6}{4} + \frac{1}{4} - \frac{6}{4} + \frac{9}{4}\right) h_1 h_2 \\
&= \frac{1}{64} \cdot 16 \cdot d_1 d_2 + 0 \cdot d_1 h_2 + 0 \cdot h_1 d_2 + \frac{1}{64} \cdot \frac{16}{4} \cdot h_1 h_2 = \frac{1}{4} d_1 d_2 + \frac{1}{16} h_1 h_2
\end{aligned}
\tag{4}
$$

And thus, the expected correlation in this locus is

$$
Cor(Y_1, Y_2) = \frac{Cov(Y_1, Y_2)}{\sqrt{Var(Y_1)}\sqrt{Var(Y_2)}} = \frac{\frac{1}{4} d_1 d_2 + \frac{1}{16} h_1 h_2}{\sqrt{\frac{1}{2} d_1^2 + \frac{1}{4} h_1^2}\sqrt{\frac{1}{2} d_2^2 + \frac{1}{4} h_2^2}} \tag{5}
$$

We may investigate how the correlation will depend on dominance deviations by varying the $h$'s between $+$ and $-$ the $d$'s. In **Supplementary Figure 8** we have plotted expected correlations over varying degrees of dominance deviations in the two genotypic effect for the different phenotypes, the $d$'s have value 1. An assumption is that $d_1$ and $d_2$ have same sign, thus the correlation is positive.

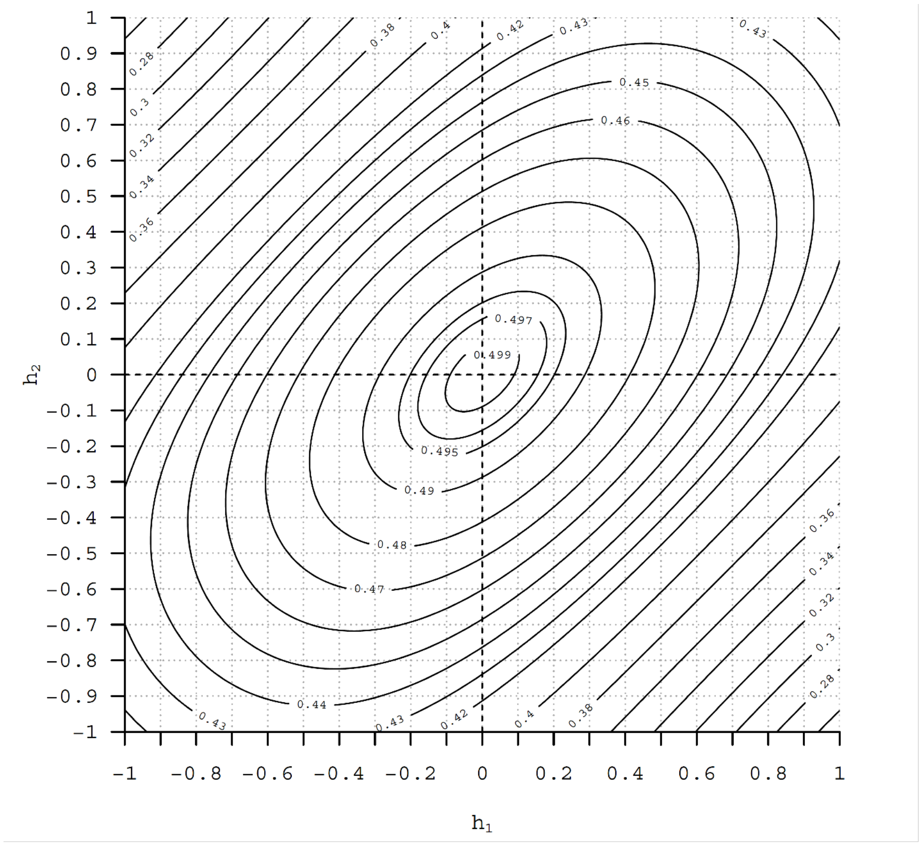

**Supplementary Figure 8.** Contour plot of deviations from additivity, when $h$'s are at $\pm 1$ there is maximal dominance deviation.

From the derivation and plot we may notice several things: 1. When one genotypic effect is at maximum dominance, and the other at perfect additivity, the correlation is $\frac{1}{\sqrt{6}} \approx 0.4082$. 2. When both genotypic effects have maximum deviation, in the same $\pm$ direction, the correlation is $\frac{5}{12} \approx 0.4167$. 3. When both genotypic effects are purely additive the correlation is $0.5$. 4. When the genotypic effects are at maximum deviation, but in opposite $\pm$ direction, the correlation is $0.25$.

Thus, the contribution to correlation between two phenotypes in different DZ twins depends on the amount and direction (i.e., dominance or recessiveness) of dominance deviation.

#### 2. The correlation depends on the allele frequencies

If we want to expand on previous simplified equal allele frequency and allow any allele frequency, we quickly generate a complex expression. Therefore, we set up a simulation with a larger number of alleles and a large number of DZ twins, where we vary the dominance deviations, to assess the impact of varying allele frequencies.

We simulated 100,000 DZ pairs and 100 assumed contributing loci under different scenarios. In all scenarios we assumed that the sign and maximal genotypic effect were the same for both phenotypes. Additional assumptions were non-assortative mating (i.e., parents were treated as two random individuals in the population), no association between minor allele frequency (MAF) and genotypic effect, no interactions between loci, and no interactions with any 'environmental' (i.e., non-genetic) variable. The genotypic effects were drawn randomly from a standard normal distribution. Each scenario is based on different MAF;

1. MAF = 0.5 for all loci.
2. MAF drawn from a uniform distribution between 0 and 0.5.
3. MAF drawn from a uniform distribution between 0 and 0.1.
4. MAF drawn from a random distribution between 0 and 0.5 with an 'L'-shape (a situation with MAF pushed towards 0 but still covering the full range).

The MAF distribution in point 4 is presented in **Supplementary Figure 9**, and was found through

$$MAF = \frac{e^{U^2} - 1}{2(e^{0.25} - 1)}, \text{where } U \sim Uniform(0, .5) \tag{6}$$

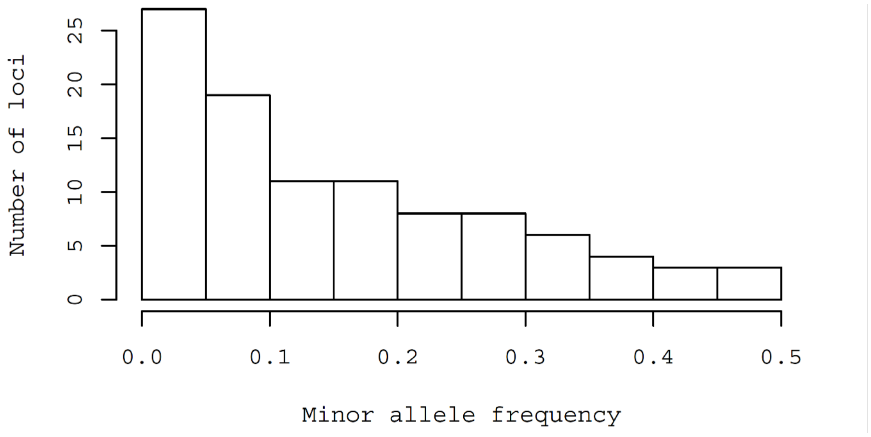

**Supplementary Figure 9.** A specific scenario for minor allele frequency distribution.

From the each of the four simulations with different MAF distributions we produce three separate series of estimates over varying dominance deviation: **scenario A**, varying degree of dominance deviation in one phenotype, and fixed additive genotypic effect in other phenotype, **scenario B**, varying degree of dominance deviation in one phenotype, and maximal dominance deviation genotypic effect in other, and **Scenario C**, varying dominance deviation vs maximal recessive deviance. Note that dominance in the *minor* allele corresponds to recessiveness in the *major* allele, and *vice versa*. These simulations serve to investigate the impact of MAF on correlation due to genetic effects between DZ twins (and full siblings) in the scenario where genotypic expression with regard to dominance deviations differ between the twins, results are presented in **Supplementary Figure 10**.

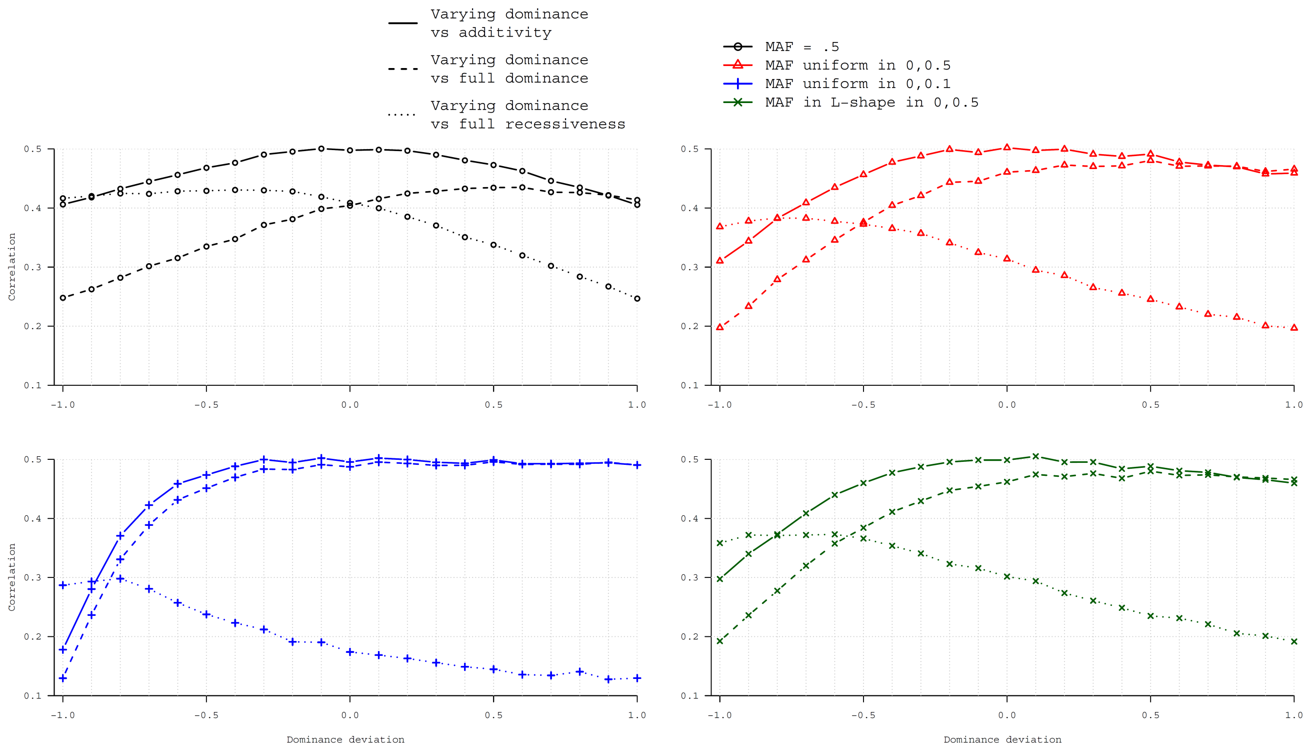

**Supplementary Figure 10.** Simulated correlation (*y-axis*) due to genetics with different MAF, and where one phenotype's genotypic effect varies in dominance deviation (the $h$ as proportion of $d$; *x-axis*) while the other's is fixed as purely additive or with maximal dominance or recessive deviation. Negative values indicate that the minor allele is recessive, positive that it is dominant.

We observe allelic frequency affect in all simulated correlations except where both individual's genotypic expression is completely additive (where the correlation between individuals is 0.5 for all MAF distributions). Further, we can see that **scenario A** has a higher correlation than **scenario B** and **C**, except for when the dominance deviation goes towards its extremes, greater/lesser than approximately $\pm 0.8$. This is most clearly seen for **Scenario B**, where the minor allele is recessive.

In reality, we would not have any way of knowing the MAF distributions for contributing loci, nor would we know the dominance deviations (which would vary between loci).

#### 3. Suggested intuitive solution

We suggest that estimation of genetic correlation based on twin data based on an intuitive solution. We have observed that correlating a phenotype with varying dominance deviation between twins will produce a resulting correlation which lies between pure additivity (i.e., correlation $= 0.5$) and maximal dominance deviation for either the major or minor allele (varying resulting correlation depending on MAF) – except for when the dominance deviations are nearing its maxima or minima. Hence, if the classic twin model correctly captures the additive genetic effects ($A$) and dominance deviations ($D$) for each trait separately, we can suggest an intuitive modeling approach to estimate the genetic correlation between the two which relies on placing the resulting correlation in between that of $A$ and $D$ alone.

*3.1 Expected genetic correlation*

To be able to assess the performance of an estimating procedure of the genetic correlation we need to know the genetic correlation if both phenotypes were expressed in one individual. Since the solution when allele frequencies are unequal quickly become unruly, we simulate this is a similar fashion as in *Section 2* above, using the same ‘L’-shaped MAF. We simulated using 100,000 individuals per each combination of values from $-1$ to 1 by steps of 0.1 – in **Supplementary Figure 11** the result is presented.

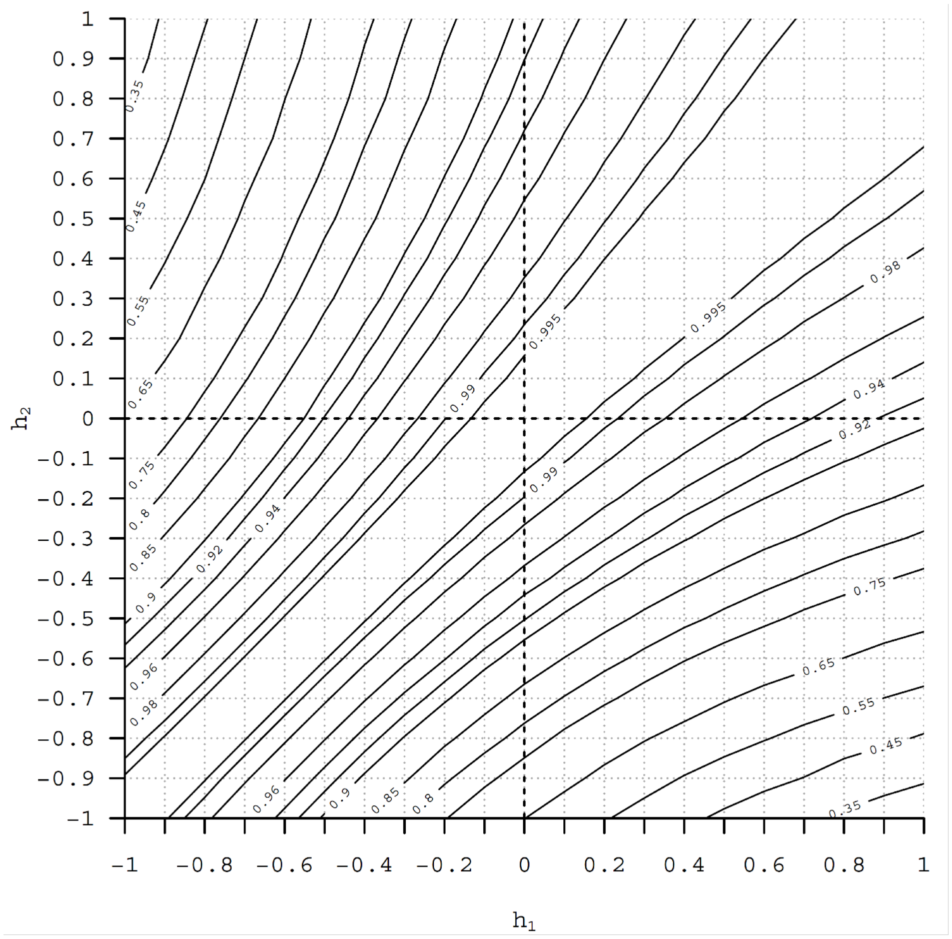

**Supplementary Figure 11.** Simulation for estimating genetic correlation. The contour plot shows resulting genetic correlation under different amounts of dominance deviation (positive) and recessive deviations (negative) for the minor allele.

We can observe that produced correlations deviate from 1 as the dominance deviations (or recessive ditto) moves away from 0 differently in the two phenotype’s genotypic effects. The genetic correlation becomes lowest when one phenotype has more dominance deviation for the minor allele and the other has more recessive deviations. We can also see that it matters which of the major or minor allele is more dominant, where the minor allele being dominant is less problematic.

*3.2 Suggested estimating procedure*

Per the classic twin model – with $A$, without $D$ and $C$ (the 'shared environment' contribution), but with $E$, the individually unique contributions to variance not shared between individuals (often referred to as an $AE$-model) – we may model the covariance between opposite sexed DZ twins (osDZ) as (see e.g. Neale and Maes [1]; here sub-index ending with $f$ indicated the female twin and sub-index ending with $m$ indicated the male twin):

$$Cov\left(\begin{bmatrix} Y_1 \\ Y_2 \end{bmatrix}\right)_{osDZ} = \begin{bmatrix} \sigma_{Af}^2 + \sigma_{Ef}^2 & r_{fm}0.5\sigma_{Af}\sigma_{Am} \\ r_{fm}0.5\sigma_{Af}\sigma_{Am} & \sigma_{Am}^2 + \sigma_{Em}^2 \end{bmatrix} \quad (7)$$

The model fitting would estimate the $r_{fm}$ as the genetic correlation between males and females, appropriately bounded between $-1$ and $1$. A direct translation into situation where the model is an $ADE$ model is

$$Cov\left(\begin{bmatrix} Y_1 \\ Y_2 \end{bmatrix}\right)_{osDZ} = \begin{bmatrix} \sigma_{Af}^2 + \sigma_{Df}^2 + \sigma_{Ef}^2 & r_{fm}0.5\sigma_{Af}\sigma_{Am} + 0.25\sigma_{Df}\sigma_{Dm} \\ r_{fm}0.5\sigma_{Af}\sigma_{Am} + 0.25\sigma_{Df}\sigma_{Dm} & \sigma_{Am}^2 + \sigma_{Dm}^2 + \sigma_{Em}^2 \end{bmatrix} \quad (8)$$

However, in this setup the estimated genetic correlation is not allowing $D$ to contribute to estimation of $r_{fm}$. A natural expansion with the inclusion of $D$ is

$$Cov\left(\begin{bmatrix} Y_1 \\ Y_2 \end{bmatrix}\right)_{osDZ} = \begin{bmatrix} \sigma_{Af}^2 + \sigma_{Df}^2 + \sigma_{Ef}^2 & r_{fm}\left(0.5\sigma_{Af}\sigma_{Am} + 0.25\sigma_{Df}\sigma_{Dm}\right) \\ r_{fm}\left(0.5\sigma_{Af}\sigma_{Am} + 0.25\sigma_{Df}\sigma_{Dm}\right) & \sigma_{Am}^2 + \sigma_{Dm}^2 + \sigma_{Em}^2 \end{bmatrix} \quad (9)$$

The assumption implicitly encoded here is that the same subset of loci having dominance deviations in one sex's genotypic expression also has dominance deviation expression in the other sex, while additive effects in one sex does not correlate with dominance deviations in the other sex. As shown above, if we deviate from this assumption, we will have different resulting correlations due to genetic effects. As an extreme example, from a twin modelling view-point, suppose the female trait has an estimated 0 $A$ contribution according to the model and some $D$ contribution, while the male has only $A$ contribution and no $D$; the result would be that there's no way the model could ascribe any correlation between sexes to genetics (and the modelled $r_{fm}$ would take on any value between -1 and 1 with equal likelihood). This is obviously not appropriate, since the same locus could contribute in a pure additive way for one sex's phenotype and a dominant way for the other sex's phenotype. We therefore suggest that the correlation can be modelled as (note, same equation as in *Introduction*)

$$Cov\left(\begin{bmatrix} Y_1 \\ Y_2 \end{bmatrix}\right)_{osDZ} = \begin{bmatrix} \sigma_{Af}^2 + \sigma_{Df}^2 + \sigma_{Ef}^2 & r_{fm}\left(0.5\sigma_{Af}\sigma_{Am} + 0.25\sigma_{Df}\sigma_{Dm} + 0.25\sigma_{Af}\sigma_{Dm} + 0.25\sigma_{Df}\sigma_{Am}\right) \\ r_{fm}\left(0.5\sigma_{Af}\sigma_{Am} + 0.25\sigma_{Df}\sigma_{Dm} + 0.25\sigma_{Af}\sigma_{Dm} + 0.25\sigma_{Df}\sigma_{Am}\right) & \sigma_{Am}^2 + \sigma_{Dm}^2 + \sigma_{Em}^2 \end{bmatrix} \quad (1)$$

This suggestion is based on the observed simulated correlations behavior towards the extremes of the dominance deviation (**Supplementary Figure 10**), where correlation due to genetics between DZ twins seem to be most affected by differences in dominance deviations. Not allowing genetics to contribute to the correlation across individuals $A\ and\ D\ sources$, may cause bias; and, towards the extremes, the correlation between additive and dominant/recessive genetic effects resemble that of dominance-to-dominance correlation behavior (although, not an exact correspondence between the two). As such, we expect the performance of the estimating procedure to be most suitable for scenarios where the $A$ and $D$ contributions to phenotypic variance differ considerable between males and females.

To investigate the performance of this suggested modelling approach we performed a series of simulations. For each simulation we used the above approach of simulating parental alleles and drawing offspring alleles, and then we created same sexed MZ and DZ twin pairs in addition to the opposite sexed DZ pairs. We assumed 7000 pairs for each sex-zygosity combination. Additionally, we estimated the resulting genetic correlation, to be able to compare the produced correlation with. We added an individually unique variation (the $E$), drawn from a random normal distribution with the same variance as the simulated genetic variance separately by sex, and used the classic twin methodology with sex-limitation models to the data. We fitted the models using equations (1) (our suggestion), (9), (8), and (7), to provide an overall picture of the performance of different approaches. We **a.** investigated the full range of dominance deviations (all combinations of the $h$'s between $-1$ and 1 by 0.25 steps), **b.** investigated the situation where one phenotype is additive and the other has dominance deviations (the $h_1$ in $-1.0$ to $-0.8$ and 0.8 to 1.0 in 0.05 steps, the $h_2$ fixed at 0), and **c.** did a more thorough investigation on the extreme values (the $h_1$ in $-1.0$ to $-0.8$ and $h_2$ in 1.0 to 0.8 in 0.05 steps) – for each combination the simulation was run 10 times. In the simulation we used the same 'L'-shaped MAF as above. In **Supplementary Figure 12a**, **12b**, and **12c** the results of these three sets of simulations are shown. We plot the difference between estimated $r_{fm}$ and calculated true $r_G$ as an estimate of bias in the separate estimating approaches.

**a.** Full range of dominance deviations

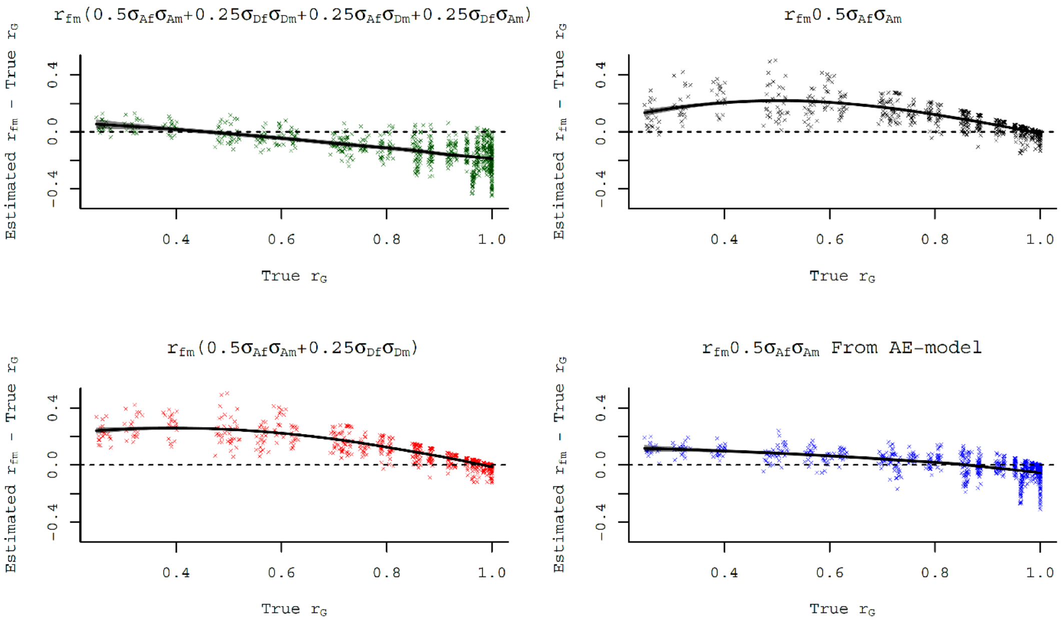

**b.** Dominance deviations versus additive

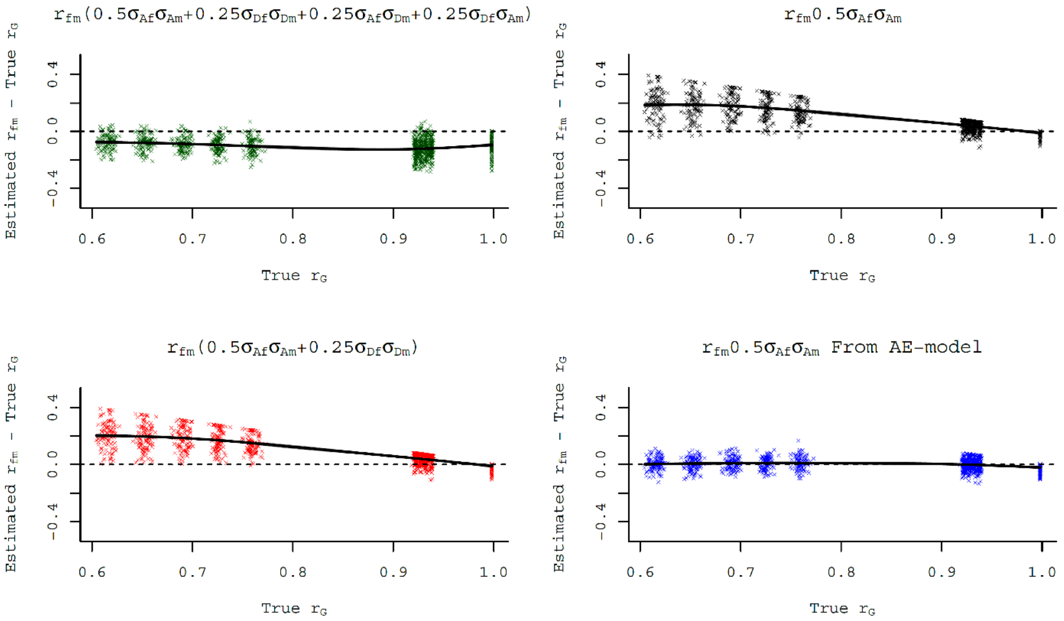

**c.** Dominance deviations towards extreme

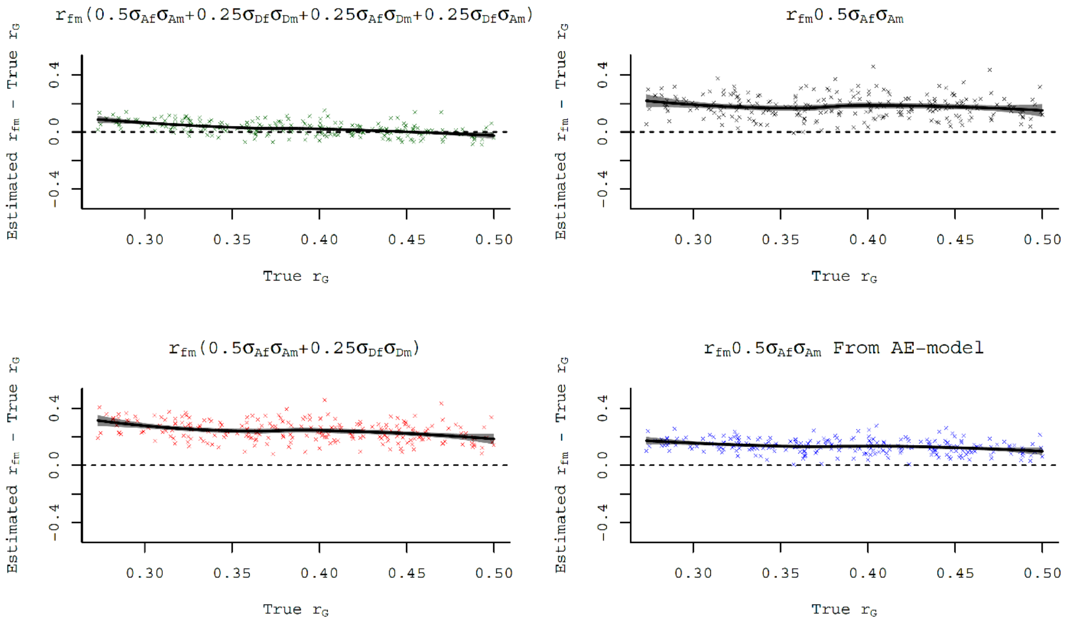

**Supplementary Figure 12.** Performance of different estimation procedures in a simulation. A locally smoothed polynomial regression line is fitted to each scenario (using 'loess' function in R).

We summarize the behavior of the four different estimating approaches in **Supplementary Table 9**. We observe the following features of the four different estimating approaches:

1. Neither estimating approach has as good performance throughout the dominance deviation range.
2. Our suggested estimating approach has best performance in the most extreme scenarios.
3. Our suggested estimating approach has a negative bias for most scenarios, and only slightly positive bias when dominance deviations are towards the very extremes.
4. All standard estimating approach (i.e., the three simulated estimating approach except our suggestion) generally has a upwards bias, towards 1.0.
5. The modelling approach of the $AE$-model perform better than other estimating approach, except for the very extremes, with regard to bias (low mean squared error) but precision may be overestimated, i.e. too low standard errors (with relatively lower coverage probability compared with standard approaches).

**Supplementary Table 9.** Performance over different scenarios for the investigated estimating procedures

| | **a.** Full range of dominance deviations | **b.** Dominance deviations versus additive | **c.** Dominance deviations towards extreme |
|---|---|---|---|
| Mean squared error | | | |
| $r_{fm}(0.5\sigma_{Af}\sigma_{Am} + 0.25\sigma_{Df}\sigma_{Dm} + 0.25\sigma_{Af}\sigma_{Dm} + 0.25\sigma_{Df}\sigma_{Am})$ | 0.029 | 0.015 | 0.003 |
| $r_{fm}0.5\sigma_{Af}\sigma_{Am} + 0.25\sigma_{Df}\sigma_{Dm}$ | 0.016 | 0.020 | 0.038 |
| $r_{fm}(0.5\sigma_{Af}\sigma_{Am} + 0.25\sigma_{Df}\sigma_{Dm})$ | 0.020 | 0.021 | 0.063 |
| $r_{fm}0.5\sigma_{Af}\sigma_{Am}$ from $AE$-model | 0.007 | 0.002 | 0.021 |
| Mean error | | | |
| $r_{fm}(0.5\sigma_{Af}\sigma_{Am} + 0.25\sigma_{Df}\sigma_{Dm} + 0.25\sigma_{Af}\sigma_{Dm} + 0.25\sigma_{Df}\sigma_{Am})$ | -0.126 | -0.103 | 0.024 |
| $r_{fm}0.5\sigma_{Af}\sigma_{Am} + 0.25\sigma_{Df}\sigma_{Dm}$ | 0.074 | 0.100 | 0.178 |
| $r_{fm}(0.5\sigma_{Af}\sigma_{Am} + 0.25\sigma_{Df}\sigma_{Dm})$ | 0.086 | 0.105 | 0.242 |
| $r_{fm}0.5\sigma_{Af}\sigma_{Am}$ from $AE$-model | -0.001 | 0.001 | 0.134 |
| Proportion negative bias | | | |
| $r_{fm}(0.5\sigma_{Af}\sigma_{Am} + 0.25\sigma_{Df}\sigma_{Dm} + 0.25\sigma_{Af}\sigma_{Dm} + 0.25\sigma_{Df}\sigma_{Am})$ | 0.872 | 0.935 | 0.318 |
| $r_{fm}0.5\sigma_{Af}\sigma_{Am} + 0.25\sigma_{Df}\sigma_{Dm}$ | 0.234 | 0.112 | 0.008 |
| $r_{fm}(0.5\sigma_{Af}\sigma_{Am} + 0.25\sigma_{Df}\sigma_{Dm})$ | 0.229 | 0.096 | 0.000 |
| $r_{fm}0.5\sigma_{Af}\sigma_{Am}$ from $AE$-model | 0.499 | 0.463 | 0.000 |
| Coverage probability | | | |
| $r_{fm}(0.5\sigma_{Af}\sigma_{Am} + 0.25\sigma_{Df}\sigma_{Dm} + 0.25\sigma_{Af}\sigma_{Dm} + 0.25\sigma_{Df}\sigma_{Am})$ | 0.602 | 0.721 | 0.895 |
| $r_{fm}0.5\sigma_{Af}\sigma_{Am} + 0.25\sigma_{Df}\sigma_{Dm}$ | 0.862 | 0.895 | 0.448 |
| $r_{fm}(0.5\sigma_{Af}\sigma_{Am} + 0.25\sigma_{Df}\sigma_{Dm})$ | 0.755 | 0.841 | 0.054 |
| $r_{fm}0.5\sigma_{Af}\sigma_{Am}$ from $AE$-model | 0.768 | 0.965 | 0.192 |

##### *3.3 Conclusion*

In a scenario with diverging dominance and additive contributions to phenotypic variance in males and females using the classic twin model, the standard estimating approaches has the feature that they overestimate the genetic correlation between males and females (which we here call $r_{fm}$). Thus, even if there is a genetic correlation lower than 1.0 the estimating approaches will have a lower likelihood of detecting it since they are biased upwards. In contrast, using our suggested approach will produce an estimate which, if it is biased at all, will be biased downwards. Therefore, if a model fitted with our estimating approach produces a $r_{fm}$ which is not statistically significant, it is likely a true null finding. Additionally, if fitting an $AE$-model is not appropriate (e.g., due to poor model fit), our suggested estimating approach has a lower mean squared error than standard estimating approach in extreme scenarios (with a larger proportion of negative bias).
